# Characterization and Validation of Adverse Childhood Experiences Data in the All of Us Research Program

**DOI:** 10.64898/2026.07.25.26358793

**Authors:** Daniel Musachio, Camille Settles, Suzi Hong, Kit Curtius, William Perry, Colin G. Walsh, Amy M. Sitapati

**Affiliations:** Division of Biomedical Informatics, The University of California, San Diego, Biomedical Research Facility II (BRF2), 5A, 9500 Gilman Drive, La Jolla, California 92037, USA; Jacobs School of Engineering, The University of California, San Diego, 9500 Gilman Drive, La Jolla, California 92037, USA; Department of Statistics and Applied Probability, The University of California, Santa Barbara, Santa Barbara, California 93106, USA; Herbert Wertheim School of Public Health and Human Longevity Science, 9500 Gilman Drive, La Jolla, California 92037, USA; VA San Diego Healthcare System, San Diego, California 92161, USA; Moores Cancer Center, University of California, San Diego, California 92037, USA; Department of Psychiatry, The University of California, San Diego, 200 West Arbor Dr, San Diego, California 92103, USA; Department of Biomedical Informatics, Vanderbilt University Medical Center, 2525 West End Ave, Suite 1475, Nashville, Tennessee 37203, USA; Department of Medicine, Vanderbilt University Medical Center, 2525 West End Ave, Suite 1475, Nashville, Tennessee 37203, USA; Department of Psychiatry and Behavioral Sciences, Vanderbilt University Medical Center, 2525 West End Ave, Suite 1475, Nashville, Tennessee 37203, USA; Institute for Applied Health Intelligence, The University of California San Diego, 9500 Gilman Drive, La Jolla, California 92037, USA

**Keywords:** Adverse childhood experiences, All of Us Research Program, Childhood adversity, Precision medicine, Social determinants of health, Epidemiology

## Abstract

Adverse childhood experiences (ACEs) are major determinants of lifelong health, yet few large precision medicine cohorts integrate standardized ACE measures with longitudinal clinical, genomic, and participant-reported data. In this cross-sectional study, we characterized the newly released 11-item ACE questionnaire using data from 137,946 All of Us Emotional Health History and Well-Being survey respondents. 90,540 completed all 11 items and 91,871 could be classified across all eight Centers for Disease Control and Prevention (CDC) ACE domains. The questionnaire demonstrated good internal consistency (Kuder-Richardson Formula 20 = 0.79), and the derived eight-domain score showed good reliability (Kuder-Richardson Formula 20 = 0.75). Compared with participants eligible to complete the survey, respondents were disproportionately White and non-Hispanic, whereas Black or African American and Hispanic participants were underrepresented. Increasing ACE burden was independently associated with higher odds of clinical and social determinant outcomes, with the strongest associations observed for post-traumatic stress disorder, food insecurity, bipolar disorder, suicidal ideation and self-harm, and substance use disorder. Outcome prevalence generally increased with ACE burden, supporting dose-response relationships. These findings establish the All of Us ACE dataset as a reliable resource for epidemiologic, clinical, genomic, and precision medicine research on childhood adversity.

## 1. Introduction

Adverse childhood experiences (ACEs) are potentially traumatic events occurring before 18 years of age, including abuse, neglect, and household dysfunction, that are strongly associated with adverse physical, mental, and social outcomes throughout individual lifespan [1–3]. Higher cumulative ACE burden has consistently been linked to increased risk of psychiatric disorders, substance use disorders, chronic medical conditions, and premature mortality across diverse populations [1,4,5]. Consequently, ACEs are recognized as a major contributor to adverse health outcomes over the lifespan and have become important targets for prevention and early intervention, which underscores their significance in precision medicine research and clinical care [1,2].

Despite decades of research, important gaps remain in understanding the biological and clinical mechanisms through which childhood adversity influences lifelong health [1]. Although numerous cohort studies have advanced ACE research, many are limited by insufficient demographic diversity, predominantly cross-sectional data, or limited integration with longitudinal health information [2,6]. For example, although the UK Biobank helps uncover the health impact of childhood adversity data through the five-item Childhood Trauma Screener, its analytic cohorts remain racially homogeneous composed predominantly of White participants (97.2% in one recent ACE study) [7] and primarily include middle-aged and older adults, limiting the generalizability of findings to broader and more diverse populations. Furthermore, its childhood adversity assessments are limited to five abuse and neglect domains rather than the broader spectrum of adverse childhood experiences assessed in other ACE studies [6,7]. These limitations highlight the need for large, diverse longitudinal cohorts that integrate standardized ACE measures with multimodal health data to better understand the biological, behavioral, and environmental pathways linking childhood adversity to health over extended lifespan [8]. The All of Us Research Program was established to accelerate precision medicine by creating one of the largest and most diverse biomedical research cohorts in the United States, integrating electronic health records, participant surveys, biospecimens, genomic data, physical measurements, and longitudinal digital health information [8,9]. Through intentional recruitment of populations historically underrepresented in biomedical research, All of Us provides a unique opportunity to investigate health disparities and improve the generalizability of scientific findings [8,9].

In June of 2026, the All of Us Research Program released the 11-item Adverse Childhood Experiences questionnaire data within the Emotional Health History and Well-Being survey, providing a valuable opportunity to investigate childhood adversity in a large, deeply phenotyped U.S. cohort [10]. However, the completeness, representativeness, and measurement properties of these newly released data have not yet been systematically characterized, limiting their broader application in epidemiologic, clinical, and translational research. Here, we characterize and evaluate the All of Us ACE survey by assessing survey completeness, demographic representativeness, internal consistency, ACE burden, convergent validity with established clinical and social outcomes, and dose-response relationships. By establishing the completeness, reliability, and validity of these newly released data, this work provides the methodological foundation necessary to support rigorous investigations of childhood adversity within one of the largest and most diverse U.S. biomedical research cohorts, enabling future epidemiologic, clinical, genomic, and precision medicine studies.

## 2. Results

### 2.1. Cohort Assembly, Survey Completion, and Representativeness

A total of 747,029 participants were included in the All of Us Controlled Tier (CDRv9) *survey_conduct* table, of whom 670,930 were eligible to complete the Emotional Health History and Well-Being (EHHWB) survey after completing the three required baseline surveys (The Basics, Overall Health, and Lifestyle) [11]. Among eligible participants, 137,913 (20.6%) completed the EHHWB survey. Thirty-three participants completed the EHHWB without record of the prerequisite surveys. Combining both groups of respondents, 137,498 had at least one valid (non-skip, non-null) response to an Adverse Childhood Experiences (ACE) questionnaire item, 90,540 completed all 11 ACE questionnaire items, and 91,871 had sufficient information to classify all eight ACE domains, which were derived by collapsing the 11 questionnaire items into eight standardized domains (e.g., the three sexual abuse items were combined into a single domain), and were included in analyses of the eight-domain ACE burden score (Figure 1).

**Figure 1.**
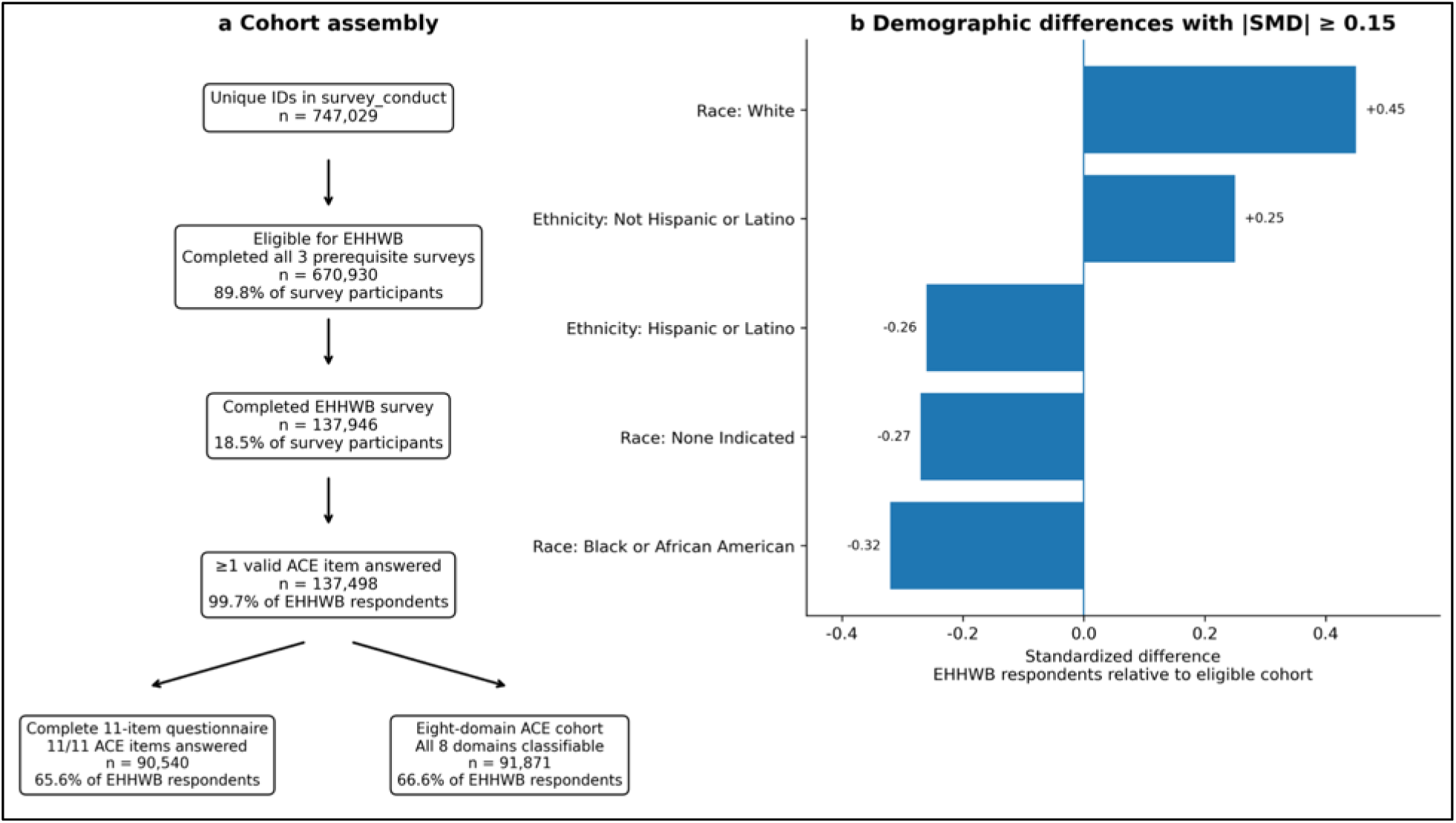
Cohort assembly yielded the final ACE analytic cohorts and revealed demographic differences in EHHWB survey response. **(a)** Flow diagram illustrating cohort assembly from the All of Us Controlled Tier (CDRv9) survey population through participants eligible to complete the Emotional Health History and Well-Being (EHHWB) survey and EHHWB survey respondents, followed by the analytic cohorts with at least one valid Adverse Childhood Experiences (ACE) questionnaire response, complete responses to all 11 ACE questionnaire items, and sufficient information to classify all eight ACE domains. Thirty-three EHHWB respondents who had not completed all three prerequisite surveys were excluded from eligibility-based demographic analyses but retained in ACE burden analyses. **(b)** Demographic differences between EHHWB survey respondents eligible to complete the EHHWB survey and all participants eligible to complete the EHHWB survey, displayed as standardized mean differences (|SMD| ≥ 0.15).

Of the 137,946 EHHWB respondents, 47,406 (34.37%) had one or more skipped or missing responses. The distribution of participants with 0–11 valid ACE questionnaire responses is presented in Supplementary Table S1. Across individual ACE questionnaire items, valid response rates ranged from 88.97% to 98.58%, with skipped response rates ranging from 0.66% to 5.33% and missing (NULL) response rates ranging from 0.76% to 10.21% (Supplementary Table S2). Questionnaire completeness differed across demographic groups (Supplementary Table S3), with Black or African American participants completing an average of 9.94 ACE questionnaire items and Hispanic or Latino participants completing 10.07 items, compared with 10.37 items among White participants and 10.33 items among participants who were not Hispanic or Latino.

Comparison of EHHWB respondents with participants eligible to complete the EHHWB survey revealed differences in demographic composition (Table 1). Meaningful differences were defined as an absolute standardized mean difference (|SMD|) ≥ 0.15. White and non-Hispanic participants were overrepresented among EHHWB survey respondents (absolute SMDs = 0.45 and 0.25, respectively), whereas Black or African American and Hispanic participants were underrepresented (absolute SMDs = 0.32 and 0.26, respectively). Respondents also exhibited an older birth-year distribution than the eligible population (median birth year, 1963 vs. 1968), with greater representation of earlier birth cohorts (Supplementary Figure S1), while gender differences were comparatively modest.

**Table 1.**
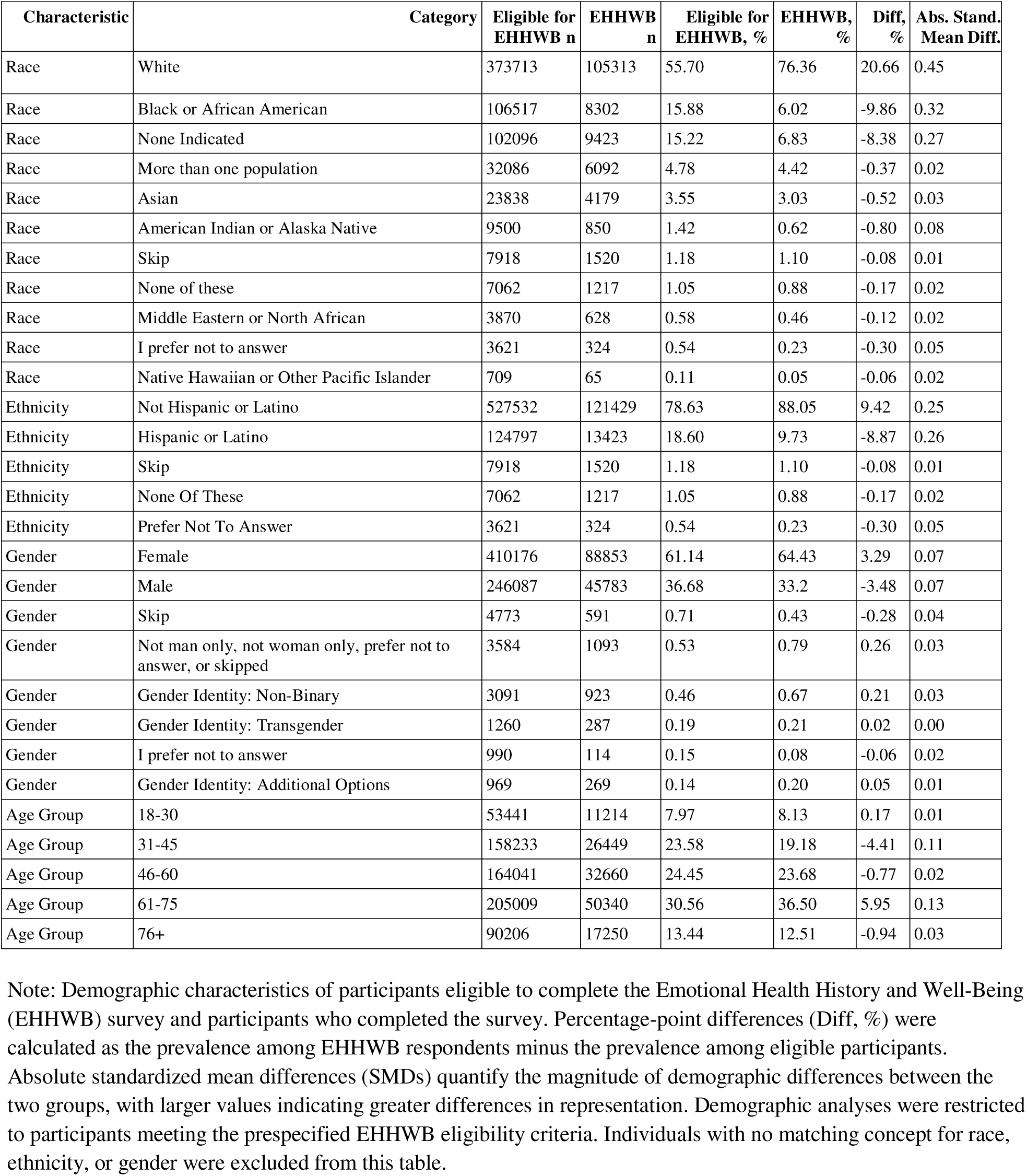
Demographic characteristics of participants eligible to complete the Emotional Health History and Well-Being (EHHWB) survey and EHHWB survey respondents.

### 2.2. Distribution of ACE Burden

Among the 91,871 participants included in the eight-domain ACE burden analyses, the mean ACE burden score was 2.11 (SD 2.07), the median was 2, and the interquartile range (IQR) was 0–3, with observed scores ranging from 0 to 8 (Table 2). The overall distribution of ACE burden scores is shown in Figure 2. Consistent with prior U.S. population estimates, ACE exposure was common, with 71.46% of participants reporting at least one ACE domain and 24.11% reporting four or more domains, compared with 63.9% and 17.3%, respectively, in the CDC Behavioral Risk Factor Surveillance System [12].

**Figure 2.**
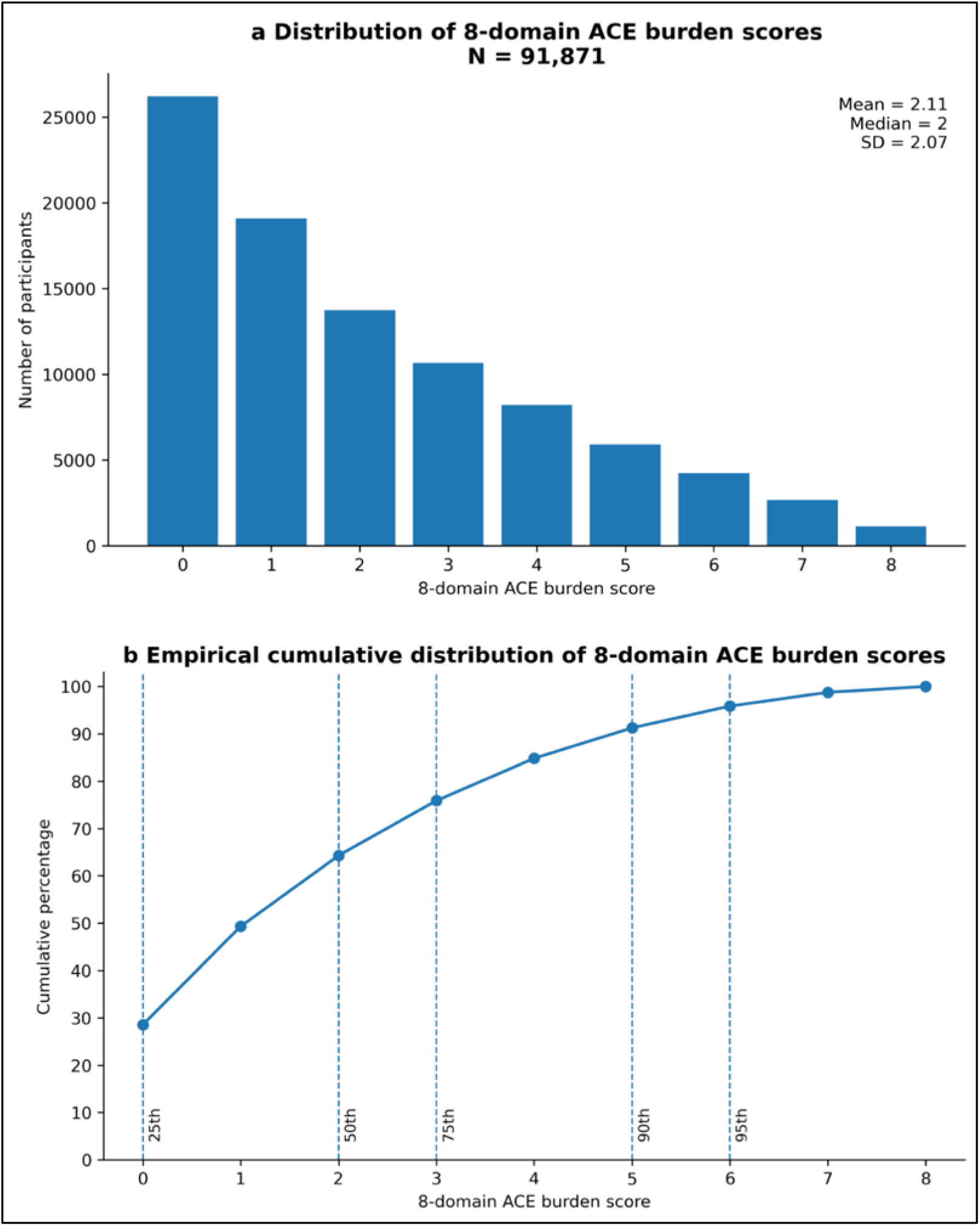
Eight-domain ACE burden scores ranged from 0 to 8 and were concentrated at lower values. **(a)** Distribution of eight-domain Adverse Childhood Experiences (ACE) burden scores among participants with sufficient information to classify all eight ACE domains (N = 91,871). Summary statistics include the mean (2.11), median (2), interquartile range (IQR, 0–3), and standard deviation (2.07). **(b)** Empirical cumulative distribution of eight-domain ACE burden scores showing the cumulative percentage of participants with ACE burden scores less than or equal to each score. Dashed lines indicate the 25th, 50th, 75th, 90th, and 95th percentiles.

**Table 2.** Distribution of eight-domain ACE burden scores among participants included in the eight-domain ACE burden analyses.

| ACE Burden Score | n | % | Cumulative n (≥ score) | Cumulative % (≥ score) |
| --- | --- | --- | --- | --- |
| 0 | 26218 | 28.54 | 91871 | 100.00 |
| 1 | 19090 | 20.78 | 65653 | 71.46 |
| 2 | 13735 | 14.95 | 46563 | 50.68 |
| 3 | 10674 | 11.62 | 32828 | 35.73 |
| 4 | 8206 | 8.93 | 22154 | 24.11 |
| 5 | 5906 | 6.43 | 13948 | 15.18 |
| 6 | 4232 | 4.61 | 8042 | 8.75 |
| 7 | 2677 | 2.91 | 3810 | 4.15 |
| 8 | 1133 | 1.23 | 1133 | 1.23 |

Note: Distribution of eight-domain Adverse Childhood Experiences (ACE) burden scores among the 91,871 participants with sufficient information to classify all eight ACE domains. The table presents the number and percentage of participants with each ACE burden score (0–8) together with the cumulative number and cumulative percentage of participants with each ACE burden score or higher ( ≥ score).

The most frequently observed ACE burden score was 0, reported by 26,218 (28.54%) participants, whereas 1,133 (1.23%) participants reported the maximum ACE burden score of 8. Overall, 46,563 (50.68%) participants reported an ACE burden score of ≥2, 22,154 (24.11%) reported a score of ≥4, and 3,810 (4.15%) reported a score of ≥7 (Table 2; Figure 2).

### 2.3. Demographic Differences in ACE

ACE burden varied substantially across demographic groups (Figure 3; Supplementary Table S5). Mean ACE burden scores were highest among Non-Binary individuals (Mean = 3.87, SD = 2.31), followed by Transgender individuals (Mean = 3.76, SD = 2.36), participants reporting additional gender identities (Mean = 3.61, SD = 2.14), multiracial participants (Mean = 3.20, SD = 2.39), and American Indian or Alaska Native participants (Mean = 3.18, SD = 2.47), whereas participants aged 76 years or older had the lowest mean ACE burden score (Mean = 1.24, SD = 1.50). Similar patterns were observed for median ACE burden scores. Overall, multiracial participants, American Indian or Alaska Native participants, Black or African American participants, Hispanic participants, and gender-diverse participants exhibited higher ACE burden than their respective reference groups (Supplementary Table S6).

**Figure 3.**
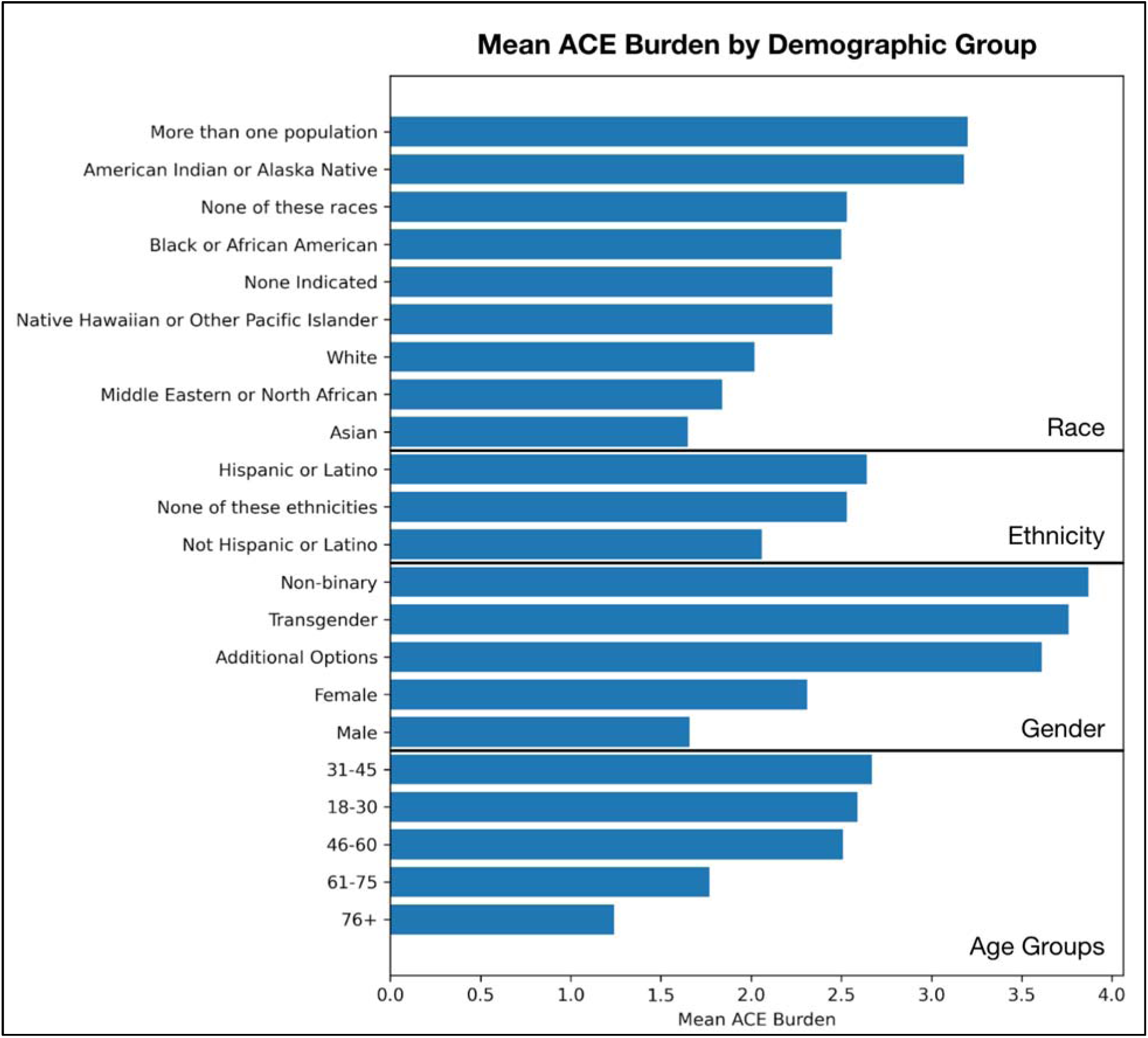
Mean eight-domain ACE burden scores varied across demographic groups. Mean eight-domain Adverse Childhood Experiences (ACE) burden score by age group, gender, race, and ethnicity among participants with sufficient information to classify all eight ACE domains (N = 91,871). Corresponding mean and median ACE burden scores are presented in Supplementary Table S5.

Multivariable logistic regression demonstrated that demographic differences in ACE burden persisted after adjustment for age group, gender, race, and ethnicity (Supplementary Table S6). Compared with their respective reference groups (White, Not Hispanic or Latino, Female, 61-75 years old), younger participants, gender-diverse participants, multiracial participants, American Indian or Alaska Native participants, Black or African American participants, and Hispanic participants had significantly greater odds of elevated ACE burden across one or more ACE thresholds. Non-Binary participants exhibited the strongest associations, with adjusted odds ratios of 3.06 (95% CI 2.44–3.83) for ACE burden ≥2, 2.32 (95% CI 1.95–2.76) for ACE burden ≥4, and 2.33 (95% CI 1.83–2.96) for ACE burden ≥7. Complete regression results are presented in Supplementary Table S6.

### 2.4. Internal Consistency

The original 11-item ACE questionnaire demonstrated good internal consistency within the All of Us cohort, with a Kuder–Richardson Formula 20 (KR-20) coefficient of 0.794, calculated after excluding non-classifiable (missing/uncertain) item responses. The KR-20 coefficient measures the internal consistency of dichotomous (yes/no) questionnaire items, with values ≥0.70 generally considered acceptable and higher values indicating greater reliability. Corrected item-total correlations ranged from 0.364 to 0.518, indicating that all questionnaire items contributed positively to the overall questionnaire score. Deletion of individual questionnaire items resulted in minimal changes to internal consistency, with KR-20 values ranging from 0.770 to 0.788 (Supplementary Table S8).

The derived eight-domain ACE burden score similarly demonstrated good internal consistency (KR-20 = 0.753), also exceeding the conventional threshold for acceptable reliability. Corrected item-total correlations ranged from 0.332 to 0.540 across the eight ACE domains, and deletion of individual domains resulted in only modest changes in reliability, with KR-20 values ranging from 0.711 to 0.748 (Supplementary Table S9).

### 2.5. Convergent Validity

Increasing eight-domain ACE burden score was associated with increased odds of all predefined clinical and social determinant outcomes (Figure 4; Supplementary Tables S10 and S11). In adjusted logistic regression models, each one-point increase in ACE burden score was associated with increased odds of post-traumatic stress disorder (adjusted OR 1.38, 95% CI 1.36–1.41), food insecurity (adjusted OR 1.37, 95% CI 1.35–1.38), bipolar disorder (adjusted OR 1.34, 95% CI 1.31–1.36), suicidal ideation and self-harm (adjusted OR 1.31, 95% CI 1.27–1.35), substance use disorder (adjusted OR 1.26, 95% CI 1.24–1.28), social isolation (adjusted OR 1.22, 95% CI 1.21–1.23), unsafe neighborhood (adjusted OR 1.20, 95% CI 1.18–1.22), major depressive disorder (adjusted OR 1.16, 95% CI 1.15–1.17), generalized anxiety disorder (adjusted OR 1.12, 95% CI 1.11–1.14), sleep disorder (adjusted OR 1.07, 95% CI 1.06–1.08), obesity (adjusted OR 1.07, 95% CI 1.06–1.08), and chronic pain (adjusted OR 1.06, 95% CI 1.05–1.06) (all *p* < 0.001).

**Figure 4.**
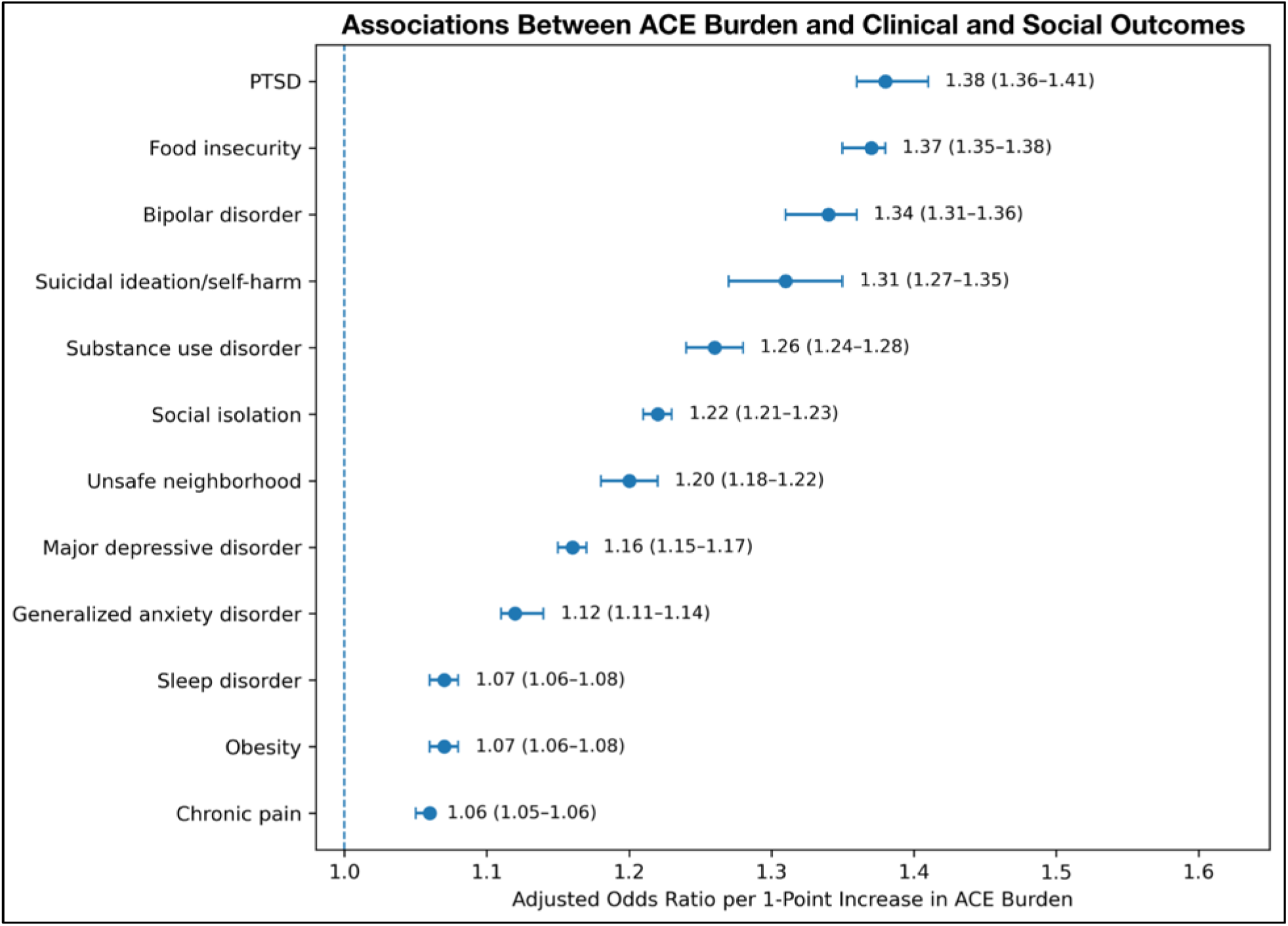
Higher eight-domain ACE burden was associated with greater odds of all predefined clinical and social determinant outcomes. Forest plot showing adjusted odds ratios (ORs) and 95% confidence intervals (CIs) for associations between the continuous eight-domain Adverse Childhood Experiences (ACE) burden score (0–8) and predefined clinical and social determinant of health (SDOH) outcomes. Odds ratios represent the change in odds associated with each one-point increase in ACE burden score after adjustment for age group, gender, race, and ethnicity.

Associations remained statistically significant after adjustment for age group, gender, race, and ethnicity, providing evidence supporting the validity of the derived eight-domain ACE burden score. Complete adjusted and unadjusted regression results are presented in Supplementary Tables S10 and S11.

### 2.6. Dose-Response Relationships

Dose-response analyses demonstrated progressively increasing prevalence for nearly all predefined clinical and social determinant outcomes with increasing eight-domain ACE burden score (Figure 5; Supplementary Figure S2; Supplementary Table S12). Across most outcomes, prevalence increased steadily with higher ACE burden, supporting a graded relationship between cumulative childhood adversity and adverse health and social outcomes.

**Figure 5.**
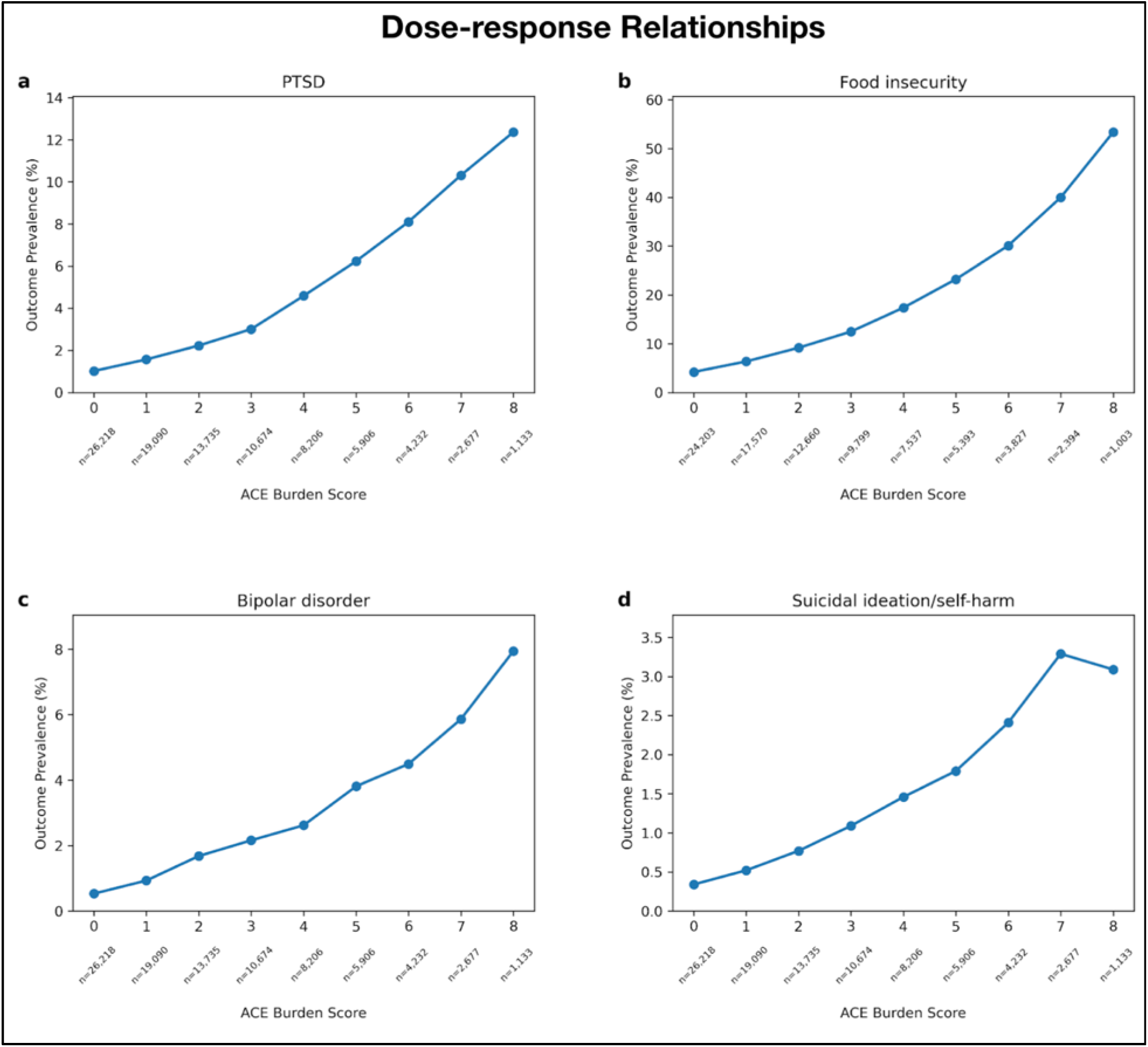
The prevalence of four representative outcomes generally increased with eight-domain ACE burden. Representative dose-response plots showing the prevalence of selected clinical and social determinant outcomes across the eight-domain Adverse Childhood Experiences (ACE) burden score (0–8). Four representative outcomes are shown: **(a)** post-traumatic stress disorder, **(b)** food insecurity, **(c)** bipolar disorder, and **(d)** suicidal ideation/self-harm; dose-response plots for the remaining eight predefined outcomes are provided in Supplementary Figure S2. Outcome prevalence was calculated separately for each ACE burden score among participants with sufficient information to classify all eight ACE domains. Sample sizes for each ACE burden score category are displayed below the x-axis.

The largest increases in outcome prevalence across the ACE burden spectrum were observed for food insecurity (4.21% at ACE score 0 to 53.34% at ACE score 8), social isolation (19.08% to 61.37%), post-traumatic stress disorder (1.02% to 12.36%), bipolar disorder (0.53% to 7.94%), suicidal ideation and self-harm (0.34% to 3.09%), and substance use disorder (2.03% to 9.62%). Major depressive disorder, generalized anxiety disorder, unsafe neighborhood, obesity, sleep disorder, and chronic pain also demonstrated increasing prevalence with higher ACE burden, although the magnitude of these associations was generally more modest (Supplementary Table S12; Supplementary Figure S2).

Small deviations from a strictly increasing pattern were observed for several outcomes at the highest ACE burden score, likely reflecting the relatively small number of participants with an ACE burden score of 8 (n = 1,133).

### 2.7. Sensitivity Analysis

In sensitivity analyses restricted to participants with at least two inpatient or outpatient visit records (N = 61,773), adjusted odds ratios remained similar to those in the primary analyses (Supplementary Tables S13–S14 and Supplementary Figure S3).

## 3. Discussion

All of Us Research Program’s recent ACEs data release offers a valuable opportunity to investigate childhood adversity within one of the largest and most diverse precision medicine cohorts worldwide [8,9]. In this initial characterization and evaluation of these newly available data, we demonstrated that the ACE questionnaire demonstrated generally high questionnaire completion and good internal consistency, that ACE burden was common among participants with sufficient information to classify all eight ACE domains, and that EHHWB respondents differed demographically from the eligible cohort, indicating evidence of selection bias. Furthermore, higher ACE burden demonstrated robust, graded associations with multiple adverse psychiatric and social determinants of health outcomes, providing evidence supporting the convergent validity of the questionnaire [13].

The All of Us Research Program substantially expands opportunities for childhood adversity research by combining standardized ACE measures with longitudinal electronic health records, biospecimens, genomic data, physical measurements, and participant-reported information in a large, diverse cohort. Unlike many previous ACE cohorts, these integrated data provide opportunities to investigate the long-term clinical, behavioral, and biological consequences of childhood adversity across the life course.

The All of Us ACE questionnaire data demonstrated strong psychometric performance, with a KR-20 coefficient of 0.79, which compares favorably with previously reported internal consistency estimates for ACE questionnaires and further supports its reliability as a measure of cumulative childhood adversity [14]. Corrected item-total correlations were consistently moderate, indicating that each questionnaire item contributed meaningfully to the overall ACE burden score. Likewise, deletion of individual items produced minimal changes in the overall KR-20 coefficient, suggesting that no single item disproportionately influenced questionnaire reliability. The derived eight-domain ACE burden score also demonstrated good internal consistency (KR-20 = 0.75), indicating that the aggregated domain-based measure retained acceptable reliability for subsequent analyses. Together, these findings indicate that the questionnaire measures a coherent underlying construct of cumulative childhood adversity in the All of Us study population and support the use of the derived eight-domain ACE burden score as a reliable proxy for cumulative ACE exposure in subsequent analyses.

ACE burden, as measured using the derived eight-domain score, also varied substantially across demographic groups, with higher levels observed among women, Black or African American participants, American Indian or Alaska Native participants, multiracial participants, Hispanic participants, and gender-diverse individuals. These findings are consistent with previous studies demonstrating that childhood adversity is not experienced uniformly across populations but instead reflects broader social, historical, and structural inequities that influence exposure to adversity early in life [1,15]. For example, greater ACEs among American Indian and Alaska Native populations may reflect the cumulative inter-generational impact of historical trauma and enduring structural inequities that influence health, health care access, and social opportunity, whereas higher ACE burden among gender-diverse individuals has been associated with minority stress, discrimination, family rejection, and other forms of social marginalization that increase vulnerability to childhood adversity [16–18]. Similarly, higher ACE burden among Black, Hispanic, and multiracial participants is consistent with previous research demonstrating that adverse childhood experiences are disproportionately distributed across historically marginalized populations and are shaped by the intersecting influences of socioeconomic disadvantage, structural inequities, and other social determinants of health [19,20]. In contrast, participants in older age groups reported lower ACE burden, which was also reported in the BRFSS [12]. This pattern may reflect birth cohort differences and changes in childhood social and environmental conditions across generations, which have shaped exposure to adverse childhood experiences over the life course and survivorship bias for those still living [13]. Together, these findings emphasize the importance of considering demographic context when interpreting ACE burden and reinforce the need to account for these factors in future epidemiologic and precision medicine studies.

The observed associations between ACE burden and adverse psychiatric, physical, and social outcomes were highly consistent with the extensive literature demonstrating that the cumulative burden of adverse childhood experiences is associated with progressively worse health outcomes across multiple domains throughout the life course [1,3,5,13]. The strongest associations were observed for post-traumatic stress disorder, food insecurity, bipolar disorder, suicidal ideation and self-harm, and substance use disorder, consistent with previous studies demonstrating that these psychiatric and social outcomes exhibit some of the strongest associations with cumulative childhood adversity [1,21–24]. Across nearly all predefined outcomes, increasing ACE burden was associated with progressively higher outcome prevalence, supporting a graded dose– response relationship rather than the existence of a discrete threshold above which risk abruptly increases.

Although completion of the ACE questionnaire was high among EHHWB respondents, participants who completed the EHHWB survey differed demographically from those who were eligible to complete the survey, with White and non-Hispanic participants overrepresented and Black or African American and Hispanic participants underrepresented. These differences likely reflect multiple sources of selection disparities, including responder bias, differential mental health survey participation, barriers to survey completion, structural differences in digital access, health literacy, and varying levels of trust and engagement with biomedical research across populations of differing race [6,25]. Such selection biases have been described across numerous large volunteer-based observational cohorts and should be considered when interpreting prevalence estimates derived from the EHHWB survey [6]. Importantly, however, these demographic differences do not diminish the substantial diversity of the All of Us cohort, which was intentionally designed to over-recruit participants from populations historically underrepresented in biomedical research and remains substantially more diverse than many commonly used biomedical research cohorts [6,8]. Furthermore, although selection bias may influence estimates of ACE prevalence within the study population, large volunteer-based cohorts can still provide informative exposure–outcome associations, provided potential sources of selection bias and the generalizability of findings are carefully considered [6,26]. Accordingly, future studies using the All of Us ACE dataset should consider these demographic differences when estimating population prevalence while recognizing the considerable strengths of the cohort for investigating exposure-outcome relationships.

An additional consideration is that ACE questionnaire completeness also varied across demographic groups, with Black or African American and Hispanic participants completing fewer questionnaire items on average than White and non-Hispanic participants. Because several groups with lower questionnaire completeness also exhibited higher ACE burden, complete-case analyses may disproportionately exclude individuals at greatest risk of childhood adversity and its downstream health consequences. This potential double disparity could result in conservative estimates of both ACE prevalence and ACE-associated health outcomes and should be considered when interpreting future analyses using these data.

This study has several important strengths. To our knowledge, it represents the first comprehensive characterization and evaluation of the newly released ACE questionnaire data from the All of Us Research Program and includes the largest characterized ACE cohort currently available within a national precision medicine resource. In addition, the use of standardized outcome definitions, prespecified analyses, and adjustment for key demographic variables strengthens the robustness and reproducibility of our findings. Nevertheless, several limitations should be considered. First, the cross-sectional design precludes causal inference and does not establish the temporal relationships between retrospectively reported ACEs and documented health outcomes, as diagnoses could occur before or after ACE questionnaire completion. Second, clinical outcomes were defined by the presence of at least one qualifying condition or observation record and were not independently validated; therefore, some outcome misclassification is possible. Third, ACEs were assessed retrospectively through self-report and are therefore subject to recall bias and potential misclassification which is the case for all ACEs studies in adults. Fourth, analyses of the derived eight-domain ACE burden score were restricted to participants with sufficient information to classify all eight ACE domains (complete-case analysis at the domain level). Although the ACE questionnaire completion was high among EHHWB respondents, restricting analyses to participants with complete data may have introduced selection bias, particularly because questionnaire completeness differed modestly across demographic groups. Finally, although adjusted analyses accounted for major demographic characteristics, residual confounding and the demographic differences observed between EHHWB respondents and the overall All of Us cohort may still influence some findings. Future longitudinal studies integrating repeated clinical measurements, electronic health records, and genomic data will be well positioned to evaluate temporal disease trajectories, resilience, and potential biological mechanisms linking childhood adversity to lifelong health outcomes [8,13].

The availability of ACE data within the All of Us Research Program creates new opportunities to investigate childhood adversity using a deeply phenotyped, longitudinal precision medicine cohort. By integrating validated ACE measures with electronic health records, genomic data, biospecimens, physical measurements, digital health information, and participant-reported outcomes, the dataset enables investigations spanning epidemiology, genomics, precision medicine, gene–environment interactions, multimodal risk prediction, resilience, and long-term disease trajectories that have not been feasible in most previous ACE cohorts [8,13]. Future research can leverage these resources and data available in the All of Us Research Program to identify genetic and environmental modifiers of ACE-associated disease risk, characterize gene– environment interactions, develop multimodal prediction models, identify factors associated with resilience despite high childhood adversity, and define longitudinal trajectories of mental and physical health across the life course [8,13]. Such work may ultimately facilitate more accurate risk stratification, earlier identification of vulnerable individuals, and the development of targeted prevention and intervention strategies informed by precision medicine approaches. By establishing the completeness, reliability, validity, and demographic characteristics of both the original 11-item ACE questionnaire and the derived eight-domain ACE burden score of these newly released data, this study provides a foundation for future epidemiologic, clinical, genomic, translational, and precision medicine investigations into the lifelong effects of childhood adversity.

## 4. Methods

### 4.1. Study Design and Data Source

This cross-sectional study used data from the NIH-funded All of Us Research Program, a nationwide precision medicine initiative designed to accelerate biomedical research through the collection of electronic health records, participant surveys, biospecimens, genomic data, physical measurements, and digital health information from a diverse U.S. population [8]. Data were obtained from the Controlled Tier (CDRv9) through the All of Us Researcher Workbench [10]. All participants provided informed consent at enrollment, and the All of Us Research Program received central institutional review board approval [8]. This study was conducted using de-identified participant data in accordance with the All of Us Research Program Data User Code of Conduct and applicable ethical guidelines [27].

### 4.2. Study Population

The source population consisted of all participants enrolled in the All of Us Research Program Controlled Tier (CDRv9) as of July 10th, 2026 [10]. Participants became eligible to complete the Emotional Health History and Well-Being (EHHWB) survey after completing the three baseline All of Us surveys (The Basics, Overall Health, and Lifestyle), which collect self-reported demographic, health, and lifestyle information [11]. Participants who completed the EHHWB survey were eligible for inclusion in the present study [28]. For participants with multiple EHHWB survey administrations, only the earliest completed survey was retained for analysis.

Participants with at least one valid response to an ACE questionnaire item were included in analyses of survey completion and data completeness. Participants with complete responses to all 11 ACE questionnaire items were included in complete-case analyses of the original 11-item questionnaire. Analyses requiring calculation of the eight-domain ACE burden score were restricted to participants for whom all eight ACE domains could be classified as either present (1) or absent (0) according to the CDC scoring framework. Responses recorded as “Skip” or missing (NULL) were considered incomplete and were not assigned an ACE value.

Among the 137,946 EHHWB respondents, 90,540 participants had complete responses to all 11 ACE questionnaire items, whereas 91,871 participants had sufficient information to classify all eight ACE domains and were included in analyses of the eight-domain ACE burden score. Participant selection is summarized in Figure 1.

### 4.3. Adverse Childhood Experiences Assessment

Adverse childhood experiences were assessed using the 11-item ACE questionnaire developed by CDC and Kaiser Permanente and administered as part of the Emotional Health History and Well-Being (EHHWB) survey in the All of Us Research Program [12,28]. The questionnaire assesses exposure to adverse experiences occurring before 18 years of age, including abuse, neglect, household dysfunction, and related childhood adversities [3].

Each ACE domain was scored dichotomously according to the CDC ACE scoring framework [12]. The eight domains measured included emotional abuse, physical abuse, sexual abuse, household substance use, household mental illness, household domestic violence, incarcerated household member, and parental separation or divorce. From the original 11 questionnaire survey, the household alcohol misuse and household drug misuse items were combined into a single household substance use domain, and the three sexual abuse items were combined into a single sexual abuse domain. Each domain was assigned a value of 1 if the participant reported exposure to that domain and 0 otherwise. Responses of “Don’t know/Not sure,” “Skip,” “No matching concept,” missing responses, and “Parents not married” for the parental separation/divorce item were classified as incomplete and were not assigned an ACE value. Exact scoring definitions are provided in Supplementary Table S4 and follow the existing CDC ACE scoring framework [12]. The eight domain scores were summed to generate a total ACE burden score ranging from 0 to 8, with higher scores indicating greater cumulative exposure to childhood adversity.

Analyses requiring calculation of the eight-domain ACE burden score were restricted to participants with sufficient information to classify each of the eight ACE domains as either present (1) or absent (0).

### 4.4. Demographic Variables

Demographic characteristics were obtained from the All of Us *person* table and included age, self-identified gender, race, and ethnicity [29]. Age was calculated as the difference between the participant’s birth year and the year of Emotional Health History and Well-Being (EHHWB) survey completion and categorized into the following groups: ≤30, 31–45, 46–60, 61–75, and ≥76 years. Birth year was additionally grouped into decade-of-birth categories (<1940, 1940– 1949, 1950–1959, 1960–1969, 1970–1979, 1980–1989, 1990–1999, and ≥2000) for descriptive analyses and visualization. Gender, race, and ethnicity were defined using participant-reported responses collected by the All of Us Research Program and mapped to standardized OMOP concepts [30].

Demographic characteristics were summarized for the overall All of Us cohort, participants eligible to complete the Emotional Health History and Well-Being (EHHWB) survey, EHHWB survey respondents, and participants included in the eight-domain ACE burden analyses. Age group, gender, race, and ethnicity were included as covariates in adjusted logistic regression models evaluating demographic predictors of elevated ACE burden and associations between ACE burden and predefined clinical and social determinant outcomes.

### 4.5. Clinical and Social Outcome Definitions

Clinical outcomes were identified using prespecified code-based definitions applied to the *condition_occurrence* and *observation* tables in the All of Us Common Data Model. Participants were classified as having an outcome if at least one qualifying record was present in either table during the available longitudinal record. This inclusive approach was used to capture clinically documented evidence represented across both tables in the heterogeneous, multisource All of Us dataset. Outcomes included major depressive disorder, generalized anxiety disorder, post-traumatic stress disorder, bipolar disorder, suicidal ideation and self-harm, substance use disorder, chronic pain, sleep disorder, and obesity. Complete definitions are provided in Supplementary Table S7.

Social determinants of health (SDOH) outcomes were derived from participant responses to the All of Us SDOH survey [31]. Outcomes included food insecurity, social isolation, and neighborhood safety. Binary outcomes were defined according to prespecified criteria provided in Supplementary Table S7.

All outcome definitions were prespecified before statistical analyses.

### 4.6. Cohort Characterization and Data Completeness

The study cohort was characterized by summarizing the number of participants with survey records, those eligible to complete the Emotional Health History and Well-Being (EHHWB) survey, EHHWB respondents, participants with at least one valid ACE response, participants with complete responses to all 11 ACE questionnaire items, and participants for whom all eight ACE domains could be classified. Participant inclusion and cohort assembly are illustrated in Figure 1a.

Data completeness was assessed by calculating the distribution of participants according to the number of valid ACE questionnaire responses (0–11). For each ACE questionnaire item, the numbers and percentages of valid responses, skipped responses, and missing responses were summarized.

To evaluate the representativeness of the EHHWB survey cohort, demographic characteristics of EHHWB respondents were compared with those of all participants eligible to complete the EHHWB survey. Comparisons included age, gender, race, and ethnicity and were summarized using counts, percentages, and absolute standardized mean differences (|SMD|). Demographic characteristics with |SMD| ≥ 0.15 are presented in Figure 1b.

### 4.7. Distribution of ACE Burden

Among participants for whom all eight ACE domains could be classified as present or absent, the distribution of the eight-domain ACE burden score was summarized using the mean, median, standard deviation, interquartile range, range, and frequency of each score from 0 to 8. The prevalence of elevated ACE burden was additionally calculated using thresholds of ≥2, ≥4, and ≥7 to capture low, high, and very high cumulative childhood adversity, respectively.

ACE burden was summarized across demographic groups defined by age, gender, race, and ethnicity. For each demographic group, the mean and median ACE burden scores were calculated.

To evaluate demographic factors associated with elevated ACE burden, separate multivariable logistic regression models were fit for ACE burden score thresholds of ≥2, ≥4, and ≥7. Each model included age group, gender, race, and ethnicity as covariates. Adjusted odds ratios, 95% confidence intervals, and two-sided P values were reported relative to prespecified reference groups.

### 4.8. Internal Consistency

Reliability was assessed for both the original 11-item ACE questionnaire and the derived eight-domain ACE burden score. For the 11-item questionnaire, corrected item-total correlations and KR-20 coefficients [32] following deletion of each individual item were calculated to evaluate the contribution of each questionnaire item to overall internal consistency (Supplementary Table S8). For the eight-domain ACE burden score, corrected item-total correlations and KR-20 coefficients following deletion of each ACE domain were similarly calculated to evaluate the contribution of each domain to overall internal consistency (Supplementary Table S9).

### 4.9. Convergent Validity

Convergent validity of the eight-domain ACE burden score was evaluated by examining associations between the continuous ACE burden score (0–8) and predefined clinical and social determinant outcomes using logistic regression models. Separate models were fit for each outcome with ACE burden score as the primary exposure variable. For each outcome, both unadjusted and adjusted logistic regression models were estimated. Adjusted models included age group, gender, race, and ethnicity as covariates. Associations are reported as odds ratios (ORs) with 95% confidence intervals (CIs) and two-sided *P* values.

### 4.10. Dose-Response Analyses

Dose-response relationships were evaluated by calculating the prevalence of each clinical and social determinant outcome across ACE burden scores. ACE burden scores were categorized from 0 to 8.

For each ACE burden score category, the number of participants with each outcome, the total number of participants, and outcome prevalence were calculated. Dose-response patterns were examined separately for clinical and social determinant outcomes and visualized as prevalence curves across increasing ACE burden scores.

### 4.11. Sensitivity Analysis

To assess the robustness of findings among participants with greater longitudinal clinical observation, we repeated the outcome analyses in a restricted cohort of participants with sufficient information to classify all eight ACE domains and at least two inpatient or outpatient visit records in the OMOP *visit_occurrence* table. Inpatient and outpatient encounters were identified using visit concept IDs 9201 and 9202, respectively. The same outcome definitions, ACE burden exposure, covariates, and regression specifications used in the primary analyses were applied.

### 4.12. Statistical Analysis

Data extraction and cohort assembly were performed using Google BigQuery within the All of Us Researcher Workbench. Statistical analyses and figure generation were conducted using Python version 3.10. Data processing was performed using pandas and NumPy, primary logistic regression analyses using statsmodels, supplementary item-level logistic regression analyses using scikit-learn, and figures were generated using Matplotlib. Descriptive statistics were used to summarize participant characteristics, survey completion, ACE burden score distributions, and outcome prevalence. Continuous variables are presented as means with standard deviations (SDs) or medians with interquartile ranges (IQRs), as appropriate, and categorical variables as counts and percentages.

Logistic regression was used because all modeled outcomes were binary. Results are reported as odds ratios (ORs) with 95% confidence intervals (CIs). All tests were two-sided with a nominal α level of 0.05. Because the analyses were intended to characterize patterns of association across predefined outcomes rather than test a single confirmatory hypothesis, no adjustment for multiple comparisons was applied. Accordingly, *p* values are nominal and were interpreted alongside effect estimates and 95% CIs. Exact *p* values are reported except when *p* < 0.001.

### 4.13. Ethics Approval

All participants in the National Institutes of Health All of Us Research Program provided written informed consent in accordance with the All of Us Institutional Review Board and the regulations and guidance of the NIH Office for Human Research Protections. This study involved secondary analysis of de-identified data accessed through the All of Us Researcher Workbench using the Data Passport model. Under this model, authorized researchers do not require separate institutional review board review because the research is not considered human subjects research, as investigators do not interact directly with participants and all data are de-identified by the All of Us Research Program. All procedures involving human participant data were performed in accordance with relevant institutional and federal guidelines and regulations and with the Declaration of Helsinki.

### 4.14. Code Availability

All custom analytical code used to generate the cohort definitions, statistical analyses, tables, and figures is available at https://github.com/dmusachio/all-of-us-ace-characterization.

### 4.15. Use of Generative Artificial Intelligence

ChatGPT 5.5 (OpenAI) was used during manuscript preparation to assist with statistical programming, code debugging, and language refinement. All analytical code, statistical analyses, results, figures, interpretations, and final manuscript content were independently reviewed, verified, and approved by the authors.

## Supporting information

STROBE Checklist

## Data Availability

The participant-level data analyzed in this study are not publicly available because the All of Us Controlled Tier contains sensitive health and survey information governed by participant privacy and data-use requirements. Controlled Tier CDRv9 is available through the secure All of Us Researcher Workbench to researchers whose institutions have executed a Data Use and Registration Agreement and who complete identity verification, required research training, including Controlled Tier training, and Data User Code of Conduct attestation. Analyses must be conducted within the secure Researcher Workbench, and participant re-identification is prohibited. Access is administered by the All of Us Research Program rather than the authors; institutional access may take from several business days to several months. Access information is available through the All of Us Researcher Workbench.

## Acknowledgements

We gratefully acknowledge *All of Us* participants for their contributions, without whom this research would not have been possible. We also thank the National Institutes of Health’s *All of Us* Research Program for making available the participant data examined in this study.

## Author Contributions

**DM:** Conceptualization, Methodology, Software, Validation, Formal analysis, Investigation, Data curation, Writing - Original draft preparation, Writing - Review & Editing, Visualization, Project administration. **CS:** Conceptualization, Methodology, Software, Validation, Formal analysis, Investigation, Data curation, Writing - Original draft preparation, Writing - Review & Editing, Visualization. **SH:** Methodology, Formal analysis, Writing - Review & Editing. **KC:** Methodology, Formal analysis, Writing - Review & Editing, Supervision. **WP:** Methodology, Writing - Review & Editing. **CGW:** Methodology, Formal analysis, Writing - Review & Editing. **AMS:** Conceptualization, Methodology, Formal analysis, Investigation, Resources, Writing - Original draft preparation, Writing - Review & Editing, Supervision, Project administration.

## Additional Information

