## Supplementary Information for "Characterization and Validation of Adverse Childhood Experiences Data in the All of Us Research Program"

**Supplementary Table S1.** Most EHHWB respondents provided valid responses to all 11 ACE questionnaire items.

| Number of valid ACE item responses | Participants, n |
|---|---|
| 0 | 448 |
| 1 | 131 |
| 2 | 71 |
| 3 | 215 |
| 4 | 897 |
| 5 | 639 |
| 6 | 1192 |
| 7 | 2844 |
| 8 | 5645 |
| 9 | 9457 |
| 10 | 25867 |
| 11 | 90540 |
| Total | 137946 |

Note: Distribution of the number of valid responses to the 11-item Adverse Childhood Experiences (ACE) questionnaire among participants who completed the Emotional Health History and Well-Being (EHHWB) survey (N = 137,946), identified within the 747,029 participants in the All of Us Controlled Tier (CDRv9) *survey_conduct* table.

**Supplementary Table S2.** Response completeness varied across the 11 ACE questionnaire items.

| ACE Questionnaire Item | Participants (N) | Valid Responses, n | Skipped Responses, n | Missing (NULL) Responses, n | Valid Responses, % | Skipped Responses, % | Missing (NULL) Responses, % |
|---|---|---|---|---|---|---|---|
| During your first 18 years of life, how often did your parents or adults in your home ever slap, hit, kick, punch or beat each other up? | 137946 | 128543 | 1409 | 7994 | 93.18 | 1.02 | 5.80 |
| During your first 18 years of life, did you live with anyone who was depressed, mentally ill, or suicidal? | 137946 | 122730 | 1127 | 14089 | 88.97 | 0.82 | 10.21 |

| | | | | | | | |
|---|---|---|---|---|---|---|---|
| During your first 18 years of life, did you live with anyone who was a problem drinker or alcoholic? | 137946 | 130317 | 3065 | 4564 | 94.47 | 2.22 | 3.31 |
| During your first 18 years of life, did you live with anyone who used illegal street drugs or who abused prescription medications? | 137946 | 128631 | 4324 | 4991 | 93.25 | 3.13 | 3.62 |
| During your first 18 years of life, did you live with anyone who served time or was sentenced to serve time in a prison, jail, or other correctional facility? | 137946 | 128664 | 7355 | 1927 | 93.27 | 5.33 | 1.40 |
| During your first 18 years of life, how often did anyone at least 5 years older than you or an adult, ever touch you sexually? | 137946 | 130907 | 2131 | 4908 | 94.90 | 1.54 | 3.56 |
| During your first 18 years of life, how often did anyone at least 5 years older than you or an adult, try to make you touch them sexually? | 137946 | 128953 | 4019 | 4974 | 93.48 | 2.91 | 3.61 |
| During your first 18 years of life, how often did anyone at least 5 years older than you or an adult, force you to have sex? | 137946 | 128214 | 5676 | 4056 | 92.95 | 4.11 | 2.94 |
| During your first 18 years of life, were your parents separated or divorced? | 137946 | 135989 | 913 | 1044 | 98.58 | 0.66 | 0.76 |
| Before age 18, how often did a parent or adult in your home ever hit, beat, kick, or physically hurt you in any way? Do not include spanking. Would you say | 137946 | 130716 | 3761 | 3469 | 94.76 | 2.73 | 2.51 |
| During your first 18 years of life, how often did a parent or adult in your home ever swear at you, insult you, or put you down? | 137946 | 125980 | 5555 | 6411 | 91.33 | 4.03 | 4.65 |

Note: Response completeness for each of the 11 Adverse Childhood Experiences (ACE) questionnaire items among participants who completed the Emotional Health History and Well-Being (EHHWB) survey (N = 137,946). For each item, the table reports the number and percentage of participants with a valid response, a skipped response, or a missing (NULL) response.

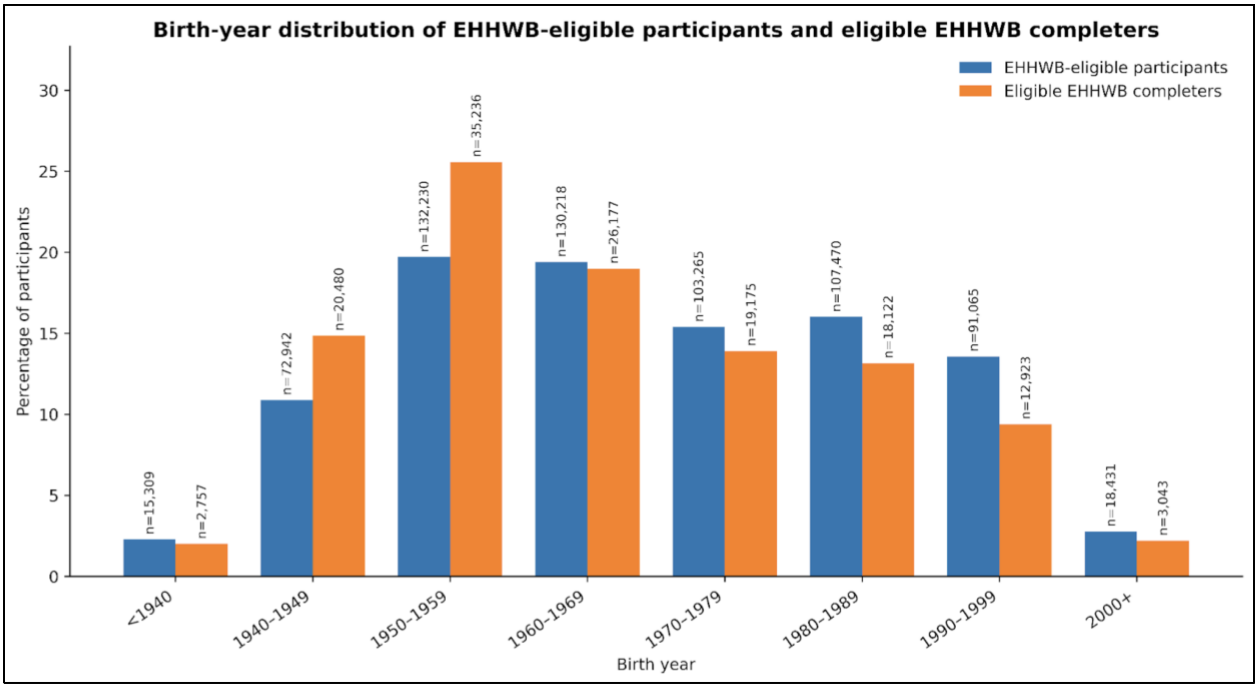


**Supplementary Figure S1.** Eligible EHHWB respondents had an earlier median birth year than the full eligible population. Distribution of participants by birth-year category among individuals eligible to complete the Emotional Health History and Well-Being (EHHWB) survey and eligible participants who completed the survey. Birth years were grouped as <1940, 1940–1949, 1950–1959, 1960–1969, 1970–1979, 1980–1989, 1990–1999, and ≥2000. Percentages were calculated within each study population using participants with available birth-year information. Eligible participants had a median birth year of 1968 (IQR, 1956–1984), whereas eligible EHHWB respondents had a median birth year of 1963 (IQR, 1953–1979). Numbers above bars indicate the number of participants in each birth-year category.

**Supplementary Table S3.** ACE questionnaire completion varied across demographic subgroups.

| Characteristic | Name | n | Mean Valid Responses | Mean Skipped Responses | Mean Missing (NULL) Response | Completed All 11 Items, % |
|---|---|---|---|---|---|---|
| Race | White | 105313 | 10.37 | 0.26 | 0.38 | 67.2 |
| Race | None Indicated | 9423 | 9.99 | 0.41 | 0.60 | 60.28 |
| Race | Black or African American | 8302 | 9.94 | 0.45 | 0.61 | 58.03 |
| Race | More than one population | 6092 | 10.21 | 0.20 | 0.59 | 60.83 |
| Race | Asian | 4179 | 10.33 | 0.26 | 0.41 | 69.16 |
| Race | Skip | 1520 | 9.76 | 0.89 | 0.35 | 57.30 |
| Race | None of these | 1217 | 10.09 | 0.31 | 0.59 | 59.57 |
| Race | American Indian or | 850 | 9.73 | 0.44 | 0.83 | 50.94 |

| | | | | | | |
|---|---|---|---|---|---|---|
| | Alaska Native | | | | | |
| Race | Middle Eastern or North African | 628 | 10.27 | 0.29 | 0.43 | 68.47 |
| Race | I prefer not to answer | 324 | 9.42 | 0.47 | 1.12 | 48.77 |
| Race | Native Hawaiian or Other Pacific Islander | 65 | 10.14 | 0.31 | 0.55 | 64.62 |
| Ethnicity | Not Hispanic or Latino | 121429 | 10.33 | 0.27 | 0.40 | 66.39 |
| Ethnicity | Hispanic or Latino | 13423 | 10.07 | 0.34 | 0.59 | 60.73 |
| Ethnicity | Skip | 1520 | 9.76 | 0.89 | 0.35 | 57.30 |
| Ethnicity | What Race Ethnicity: Race Ethnicity None Of These | 1217 | 10.09 | 0.31 | 0.59 | 59.57 |
| Ethnicity | Prefer Not To Answer | 324 | 9.42 | 0.47 | 1.12 | 48.77 |
| Gender | Female | 88853 | 10.29 | 0.28 | 0.44 | 64.98 |
| Gender | Male | 45783 | 10.32 | 0.30 | 0.38 | 67.70 |
| Gender | Not man only, not woman only, prefer not to answer, or skipped | 1093 | 10.11 | 0.09 | 0.80 | 55.90 |
| Gender | Gender Identity: Non Binary | 923 | 10.18 | 0.08 | 0.74 | 56.45 |
| Gender | Skip | 591 | 9.54 | 1.01 | 0.46 | 53.98 |
| Gender | Gender Identity: Transgender | 287 | 10.01 | 0.14 | 0.86 | 52.96 |
| Gender | Gender Identity: Additional Options | 269 | 9.95 | 0.26 | 0.78 | 52.04 |
| Gender | I prefer not to answer | 114 | 8.96 | 0.57 | 1.47 | 43.86 |
| Age Group | 18-30 | 11214 | 10.28 | 0.16 | 0.56 | 66.11 |
| Age Group | 31-45 | 26449 | 10.30 | 0.15 | 0.55 | 65.16 |
| Age Group | 46-60 | 32660 | 10.32 | 0.22 | 0.46 | 66.02 |
| Age Group | 61-75 | 50340 | 10.31 | 0.34 | 0.35 | 66.19 |
| Age Group | 76+ | 17250 | 10.17 | 0.55 | 0.28 | 63.74 |

Note: Survey completion metrics for the 11-item Adverse Childhood Experiences (ACE) questionnaire among eligible participants who completed the Emotional Health History and Well-Being (EHHWB) survey, stratified by race, ethnicity, gender, and age group. For each demographic subgroup, the table reports the number of participants, the mean number of valid responses, skipped responses, and missing (NULL) responses across the 11 ACE questionnaire items, together with the percentage of participants who completed all 11 items.

**Supplementary Table S4.** The 11 ACE questionnaire items were mapped to eight binary ACE domains.

| ACE domain | ACE questionnaire item(s) | Responses scored as 1 | Responses scored as 0 | Incomplete or no information |
|---|---|---|---|---|
| Household mental illness | During your first 18 years of life, did you live with anyone who was depressed, mentally ill, or suicidal? | Yes | No | Don't know/Not sure; Skip; No matching concept; missing response |
| Household substance use | During your first 18 years of life, did you live with anyone who was a problem drinker or alcoholic? and During your first 18 years of life, did you live with anyone who used illegal street drugs or who abused prescription medications? | Yes to either item | No to both items | No affirmative response and insufficient information to establish absence; specifically, Don't know/Not sure, Skip, No matching concept, or a missing response for at least one item when the other item is negative or also incomplete. |
| Household incarceration | During your first 18 years of life, did you live with anyone who served time or was sentenced to serve time in a prison, jail, or other correctional facility? | Yes | No | Don't know/Not sure; Skip; No matching concept; missing response |
| Parental separation or divorce | During your first 18 years of life, were your parents separated or divorced? | Yes | No | Parents not married; Don't know/Not sure; Skip; No matching concept; missing response |
| Witnessing intimate partner violence | During your first 18 years of life, how often did your parents or adults in your home ever slap, hit, kick, punch, or beat each other up? | Once; More than once | Never | Don't know/Not sure; Skip; No matching concept; missing response |
| Physical abuse | Before age 18, how often did a parent or adult in your home ever hit, beat, kick, or physically hurt you in any way? Do not include spanking. | Once; More than once | Never | Don't know/Not sure; Skip; No matching concept; missing response |
| Emotional abuse | During your first 18 years of life, how often did a parent or adult in | Once; More than once | Never | Don't know/Not sure; Skip; No matching |

| | | | | |
|---|---|---|---|---|
| | your home ever swear at you, insult you, or put you down? | | | concept; missing response |
| Sexual abuse | Before age 18, how often did anyone at least 5 years older than you or an adult: (1) touch you sexually, (2) try to make you touch them sexually, or (3) force you to have sex? | Once or More than once for any of the three items | Never for all three items | No affirmative response and insufficient information to establish absence; specifically, Don't know/Not sure, Skip, No matching concept, or a missing response for at least one item when all remaining answered items are Never or incomplete. |

Note: Each ACE domain was assigned a binary score of 0 or 1. Household alcohol misuse and household drug misuse were combined into a single household substance-use domain. The three sexual-abuse questions were combined into a single sexual-abuse domain. The eight-domain ACE burden score was calculated by summing the domain scores and ranged from 0 to 8. "Parents not married" was classified as incomplete/no information rather than scored as 0 because this response does not establish whether the participant experienced parental separation or divorce. "Don't know/Not sure," "Skip," "No matching concept," and missing responses were likewise classified as incomplete/no information. Participants with insufficient information to derive a binary (0/1) classification for one or more of the eight ACE domains were excluded from analyses of the derived eight-domain ACE burden score.

**Supplementary Table S5.** Eight-domain ACE burden scores varied across demographic subgroups.

| Characteristic | Cohort | Name | Mean ACE Burden Score | Median ACE Burden Score | Standard Deviation |
|---|---|---|---|---|---|
| Race | Complete | White | 2.02 | 1 | 2.03 |
| Race | Complete | None Indicated | 2.45 | 2 | 2.15 |
| Race | Complete | Black or African American | 2.50 | 2 | 2.15 |
| Race | Complete | More than one population | 3.20 | 3 | 2.39 |
| Race | Complete | Asian | 1.65 | 1 | 1.72 |
| Race | Complete | Skip | 1.69 | 1 | 1.86 |
| Race | Complete | None of these | 2.53 | 2 | 2.29 |
| Race | Complete | Middle Eastern or North African | 1.84 | 1 | 1.92 |
| Race | Complete | American Indian or Alaska Native | 3.18 | 3 | 2.47 |
| Race | Complete | I prefer not to answer | 2.07 | 1 | 2.18 |
| Race | Complete | Native Hawaiian or Other Pacific Islander | 2.45 | 2 | 2.19 |

| | | | | | |
|---|---|---|---|---|---|
| Ethnicity | Complete | Not Hispanic or Latino | 2.06 | 1 | 2.05 |
| Ethnicity | Complete | Hispanic or Latino | 2.64 | 2 | 2.23 |
| Ethnicity | Complete | Skip | 1.69 | 1 | 1.86 |
| Ethnicity | Complete | What Race Ethnicity: Race Ethnicity None Of These | 2.53 | 2 | 2.29 |
| Ethnicity | Complete | Prefer Not To Answer / No matching concept | 2.05 | 1 | 2.18 |
| Gender | Complete | Female | 2.31 | 2 | 2.14 |
| Gender | Complete | Male | 1.66 | 1 | 1.83 |
| Gender | Complete | Not man only, not woman only, prefer not to answer, or skipped | 3.46 | 3 | 2.21 |
| Gender | Complete | Gender Identity: Non-Binary | 3.87 | 4 | 2.31 |
| Gender | Complete | Skip / No matching concept | 1.97 | 1 | 2.04 |
| Gender | Complete | Gender Identity: Transgender | 3.76 | 4 | 2.36 |
| Gender | Complete | Gender Identity: Additional Options | 3.61 | 3 | 2.14 |
| Gender | Complete | I prefer not to answer | 3.06 | 3 | 2.33 |
| Age Group | Complete | 18-30 | 2.59 | 2 | 2.2 |
| Age Group | Complete | 31-45 | 2.67 | 2 | 2.28 |
| Age Group | Complete | 46-60 | 2.51 | 2 | 2.18 |
| Age Group | Complete | 61-75 | 1.77 | 1 | 1.85 |
| Age Group | Complete | 76+ | 1.24 | 1 | 1.50 |

Note: Mean and median eight-domain Adverse Childhood Experiences (ACE) burden scores by age group, gender, race, and ethnicity among the 91,871 participants with sufficient information to classify all eight ACE domains. The table reports the corresponding mean and median ACE burden scores. Values are reported as the mean, median, and standard deviation (SD); possible scores ranged from 0 to 8. Categories with 20 or fewer participants were omitted or combined to preserve participant privacy in accordance with All of Us reporting requirements.

**Supplementary Table S6.** Demographic characteristics were associated with elevated ACE burden at thresholds of ≥2, ≥4, and ≥7.

| Threshold | Category | Name | Reference | Model, N | Cases, N | Controls, N | Cases, % | OR (95% CI) | p-value |
|---|---|---|---|---|---|---|---|---|---|
| ACE ≥ 2 | Age Group | 31-45 | 61-75 | 91871 | 46563 | 45308 | 50.68 | 1.729 (1.664–1.796) | <0.001 |
| ACE ≥ 2 | Age Group | 46-60 | 61-75 | 91871 | 46563 | 45308 | 50.68 | 1.642 (1.586–1.700) | <0.001 |
| ACE ≥ 2 | Age Group | 76+ | 61-75 | 91871 | 46563 | 45308 | 50.68 | 0.627 (0.599–0.656) | <0.001 |

| | | | | | | | | | |
|---|---|---|---|---|---|---|---|---|---|
| ACE ≥ 2 | Age Group | <=30 | 61-75 | 91871 | 46563 | 45308 | 50.68 | 1.660 (1.573–1.753) | <0.001 |
| ACE ≥ 2 | Gender | Gender Identity: Additional Options | Female | 91871 | 46563 | 45308 | 50.68 | 2.816 (1.857–4.270) | <0.001 |
| ACE ≥ 2 | Gender | Gender Identity: Non-Binary | Female | 91871 | 46563 | 45308 | 50.68 | 3.058 (2.441–3.829) | <0.001 |
| ACE ≥ 2 | Gender | Gender Identity: Transgender | Female | 91871 | 46563 | 45308 | 50.68 | 2.577 (1.754–3.786) | <0.001 |
| ACE ≥ 2 | Gender | Male | Female | 91871 | 46563 | 45308 | 50.68 | 0.679 (0.660–0.699) | <0.001 |
| ACE ≥ 2 | Gender | Not man only, not woman only, prefer not to answer, or skipped | Female | 91871 | 46563 | 45308 | 50.68 | 2.342 (1.933–2.837) | <0.001 |
| ACE ≥ 2 | Race | American Indian or Alaska Native | White | 91871 | 46563 | 45308 | 50.68 | 1.593 (1.289–1.970) | <0.001 |
| ACE ≥ 2 | Race | Asian | White | 91871 | 46563 | 45308 | 50.68 | 0.667 (0.617–0.720) | <0.001 |
| ACE ≥ 2 | Race | Black or African American | White | 91871 | 46563 | 45308 | 50.68 | 1.336 (1.253–1.424) | <0.001 |
| ACE ≥ 2 | Race | Middle Eastern or North African | White | 91871 | 46563 | 45308 | 50.68 | 0.820 (0.677–0.993) | 0.0426 |
| ACE ≥ 2 | Race | More than one population | White | 91871 | 46563 | 45308 | 50.68 | 1.896 (1.762–2.041) | <0.001 |
| ACE ≥ 2 | Race | None Indicated | White | 91871 | 46563 | 45308 | 50.68 | 0.752 (0.678–0.834) | <0.001 |
| ACE ≥ 2 | Ethnicity | Hispanic or Latino | Not Hispanic or Latino | 91871 | 46563 | 45308 | 50.68 | 1.552 (1.419–1.698) | <0.001 |
| ACE ≥ 4 | Age Group | 31-45 | 61-75 | 91871 | 22154 | 69717 | 24.11 | 2.050 (1.964–2.141) | <0.001 |
| ACE ≥ 4 | Age Group | 46-60 | 61-75 | 91871 | 22154 | 69717 | 24.11 | 1.876 (1.801–1.953) | <0.001 |
| ACE ≥ 4 | Age Group | 76+ | 61-75 | 91871 | 22154 | 69717 | 24.11 | 0.510 (0.476–0.547) | <0.001 |
| ACE ≥ 4 | Age Group | <=30 | 61-75 | 91871 | 22154 | 69717 | 24.11 | 1.733 (1.632–1.840) | <0.001 |
| ACE ≥ 4 | Gender | Gender Identity: Additional Options | Female | 91871 | 22154 | 69717 | 24.11 | 2.073 (1.487–2.889) | <0.001 |
| ACE ≥ 4 | Gender | Gender Identity: Non-Binary | Female | 91871 | 22154 | 69717 | 24.11 | 2.318 (1.949–2.757) | <0.001 |

| | | | | | | | | | |
|---|---|---|---|---|---|---|---|---|---|
| ACE ≥ 4 | Gender | Gender Identity: Transgender | Female | 91871 | 22154 | 69717 | 24.11 | 2.681 (1.942–3.701) | <0.001 |
| ACE ≥ 4 | Gender | I prefer not to answer | Female | 91871 | 22154 | 69717 | 24.11 | 2.116 (1.196–3.743) | 0.01 |
| ACE ≥ 4 | Gender | Male | Female | 91871 | 22154 | 69717 | 24.11 | 0.620 (0.598–0.642) | <0.001 |
| ACE ≥ 4 | Gender | Not man only, not woman only, prefer not to answer, or skipped | Female | 91871 | 22154 | 69717 | 24.11 | 1.723 (1.468–2.024) | <0.001 |
| ACE ≥ 4 | Race | American Indian or Alaska Native | White | 91871 | 22154 | 69717 | 24.11 | 1.785 (1.455–2.190) | <0.001 |
| ACE ≥ 4 | Race | Asian | White | 91871 | 22154 | 69717 | 24.11 | 0.533 (0.481–0.591) | <0.001 |
| ACE ≥ 4 | Race | Black or African American | White | 91871 | 22154 | 69717 | 24.11 | 1.337 (1.248–1.433) | <0.001 |
| ACE ≥ 4 | Race | Middle Eastern or North African | White | 91871 | 22154 | 69717 | 24.11 | 0.703 (0.552–0.894) | 0.0041 |
| ACE ≥ 4 | Race | More than one population | White | 91871 | 22154 | 69717 | 24.11 | 1.958 (1.826–2.100) | <0.001 |
| ACE ≥ 4 | Race | None Indicated | White | 91871 | 22154 | 69717 | 24.11 | 0.760 (0.685–0.843) | <0.001 |
| ACE ≥ 4 | Race | None of these | White | 91871 | 22154 | 69717 | 24.11 | 1.251 (1.153–1.356) | <0.001 |
| ACE ≥ 4 | Ethnicity | Hispanic or Latino | Not Hispanic or Latino | 91871 | 22154 | 69717 | 24.11 | 1.544 (1.415–1.684) | <0.001 |
| ACE ≥ 7 | Age Group | 31-45 | 61-75 | 91871 | 3810 | 88061 | 4.15 | 3.614 (3.281–3.981) | <0.001 |
| ACE ≥ 7 | Age Group | 46-60 | 61-75 | 91871 | 3810 | 88061 | 4.15 | 2.739 (2.488–3.015) | <0.001 |
| ACE ≥ 7 | Age Group | 76+ | 61-75 | 91871 | 3810 | 88061 | 4.15 | 0.293 (0.224–0.383) | <0.001 |
| ACE ≥ 7 | Age Group | <=30 | 61-75 | 91871 | 3810 | 88061 | 4.15 | 2.874 (2.536–3.257) | <0.001 |
| ACE ≥ 7 | Gender | Gender Identity: Non-Binary | Female | 91871 | 3810 | 88061 | 4.15 | 2.331 (1.833–2.963) | <0.001 |
| ACE ≥ 7 | Gender | Gender Identity: Transgender | Female | 91871 | 3810 | 88061 | 4.15 | 2.594 (1.646–4.088) | <0.001 |
| ACE ≥ 7 | Gender | Male | Female | 91871 | 3810 | 88061 | 4.15 | 0.489 (0.447–0.535) | <0.001 |

| | | | | | | | | | |
|---|---|---|---|---|---|---|---|---|---|
| ACE ≥ 7 | Gender | Not man only, not woman only, prefer not to answer, or skipped | Female | 91871 | 3810 | 88061 | 4.15 | 1.645 (1.282–2.110) | <0.001 |
| ACE ≥ 7 | Race | American Indian or Alaska Native | White | 91871 | 3810 | 88061 | 4.15 | 2.499 (1.841–3.391) | <0.001 |
| ACE ≥ 7 | Race | Asian | White | 91871 | 3810 | 88061 | 4.15 | 0.204 (0.142–0.294) | <0.001 |
| ACE ≥ 7 | Race | Black or African American | White | 91871 | 3810 | 88061 | 4.15 | 1.188 (1.029–1.373) | 0.019 |
| ACE ≥ 7 | Race | More than one population | White | 91871 | 3810 | 88061 | 4.15 | 2.377 (2.127–2.656) | <0.001 |
| ACE ≥ 7 | Ethnicity | Hispanic or Latino | Not Hispanic or Latino | 91871 | 3810 | 88061 | 4.15 | 1.567 (1.356–1.811) | <0.001 |

Note: Multivariable logistic regression analyses evaluating demographic predictors of elevated eight-domain Adverse Childhood Experiences (ACE) burden among the 91,871 participants with sufficient information to classify all eight ACE domains. Separate models were fit for ACE burden score thresholds of ≥2, ≥4, and ≥7. Models were adjusted for age group, gender, race, and ethnicity. Results are presented as adjusted odds ratios (ORs) with 95% confidence intervals (CIs) using the prespecified reference category for each demographic variable. Administrative nonresponse categories, including skip, prefer not to answer, none of these, and no matching concept, were omitted from the table.

**Supplementary Table S7.** Clinical and social determinant outcomes were defined using electronic health record and survey data.

| Outcome | Domain | Outcome Definition | Data Source | Code Set |
|---:|---:|---:|---:|---:|
| PTSD | Clinical | ≥1 ICD-10-CM source code for PTSD | EHR condition_occurrence + observation | F43.1* |
| Food insecurity | SDOH | Positive response indicating food insecurity | Survey observation | QID 40192517; positive: 45877955, 36309834 |
| Bipolar disorder | Clinical | ≥1 ICD-10-CM source code for bipolar disorder | EHR condition_occurrence + observation | F31* |
| Suicidal ideation/self-harm | Clinical | ≥1 ICD-10-CM source code for suicidal ideation or intentional self-harm | EHR condition_occurrence + observation | R45.851; X71-X83; T14.91; T36-T65 or T71 with intentional self-harm concept name |
| Substance use disorder | Clinical | ≥1 ICD-10-CM substance use disorder source code, excluding unspecified use codes | EHR condition_occurrence + observation | F10*, F11*, F12-F16*, F18*, F19*; excludes F10.9/F10.90 etc. |

| | | | | |
|---|---|---|---|---|
| Social isolation | SDOH | Positive response indicating social isolation | Survey observation | QID 40192501; positive: 45882528, 45884455 |
| Unsafe neighborhood | SDOH | Positive response indicating disagreement that neighborhood is safe | Survey observation | QID 40192384; positive: 45884599, 45882953 |
| Major depressive disorder | Clinical | ≥1 ICD-10-CM source code for major depressive disorder | EHR condition_occurrence + observation | F32*, F33* |
| Generalized anxiety disorder | Clinical | ≥1 ICD-10-CM source code for generalized anxiety disorder | EHR condition_occurrence + observation | F41.1* |
| Sleep disorder | Clinical | ≥1 ICD-10-CM source code for sleep disorder | EHR condition_occurrence + observation | G47*, F51* |
| Obesity | Clinical | ≥1 ICD-10-CM source code for obesity | EHR condition_occurrence + observation | E66* |
| Chronic pain | Clinical | ≥1 ICD-10-CM source code for chronic pain/pain disorder | EHR condition_occurrence + observation | G89.2* |

Note: Definitions of the predefined clinical and social determinant of health (SDOH) outcomes used for convergent validity and dose-response analyses. Clinical outcomes were identified using ICD-10-CM diagnosis codes recorded in the All of Us electronic health record *condition_occurrence* and *observation* tables. SDOH outcomes were derived from participant responses to the All of Us Social Determinants of Health (SDOH) survey using prespecified questionnaire responses. Complete ICD-10-CM code sets and survey concept identifiers are provided for each outcome.

**Supplementary Table S8.** The 11-item ACE questionnaire demonstrated good internal consistency.

| Question | Positive Response, n | Total, n | Positive Response, % | Corrected item-total r | KR-20 if deleted | ΔKR-20 |
|---|---|---|---|---|---|---|
| During your first 18 years of life, did you live with anyone who was depressed, mentally ill, or suicidal? | 29432 | 90540 | 32.51 | 0.460 | 0.778 | -0.016 |
| During your first 18 years of life, did you live with anyone who was a problem drinker or alcoholic? | 24864 | 90540 | 27.46 | 0.442 | 0.779 | -0.015 |
| During your first 18 years of life, did you live with anyone who used illegal street drugs or who abused prescription medications? | 10792 | 90540 | 11.92 | 0.458 | 0.778 | -0.016 |
| During your first 18 years of life, did you live with anyone who served time or was sentenced to serve time in a prison, jail, or other correctional facility? | 5231 | 90540 | 5.78 | 0.374 | 0.787 | -0.007 |
| During your first 18 years of life, were your parents separated or divorced? | 20377 | 90540 | 22.51 | 0.364 | 0.788 | -0.006 |
| During your first 18 years of life, how often did | 18470 | 90540 | 20.40 | 0.518 | 0.770 | -0.024 |

| | | | | | | |
|---|---|---|---|---|---|---|
| your parents or adults in your home ever slap, hit, kick, punch or beat each other up? | | | | | | |
| Before age 18, how often did a parent or adult in your home ever hit, beat, kick, or physically hurt you in any way? Do not include spanking. Would you say | 22241 | 90540 | 24.56 | 0.503 | 0.772 | -0.022 |
| During your first 18 years of life, how often did a parent or adult in your home ever swear at you, insult you, or put you down? | 39900 | 90540 | 44.07 | 0.494 | 0.774 | -0.020 |
| During your first 18 years of life, how often did anyone at least 5 years older than you or an adult, ever touch you sexually? | 19334 | 90540 | 21.35 | 0.472 | 0.775 | -0.019 |
| During your first 18 years of life, how often did anyone at least 5 years older than you or an adult, try to make you touch them sexually? | 14251 | 90540 | 15.74 | 0.484 | 0.775 | -0.019 |
| During your first 18 years of life, how often did anyone at least 5 years older than you or an adult, force you to have sex? | 7419 | 90540 | 8.19 | 0.459 | 0.780 | -0.014 |

Note: Internal consistency analysis of the 11-item Adverse Childhood Experiences (ACE) questionnaire among participants with complete ACE data (N = 90,540). Analyses were restricted to participants with a positive or negative classification for all 11 questionnaire items; participants with missing, NULL, or skipped responses for any ACE item were excluded. The overall Kuder–Richardson Formula 20 (KR-20) coefficient was 0.794. For each questionnaire item, the table reports the number and percentage of participants with a positive response, the corrected item-total correlation, the KR-20 coefficient following deletion of the item, and the corresponding change in KR-20 (ΔKR-20) relative to the full questionnaire.

**Supplementary Table S9.** The derived eight-domain ACE burden score demonstrated good internal consistency.

| Domain | Positive Response, n | Total, n | Positive Response, % | Corrected item-total r | KR-20 if deleted | ΔKR-20 |
|---|---|---|---|---|---|---|
| Household mental illness | 30832 | 91871 | 33.56 | 0.477 | 0.722 | -0.031 |
| Household substance use | 29297 | 91871 | 31.89 | 0.473 | 0.723 | -0.030 |
| Incarcerated household member | 5217 | 91871 | 5.68 | 0.332 | 0.748 | -0.005 |
| Parental separation or divorce | 22006 | 91871 | 23.95 | 0.392 | 0.738 | -0.015 |
| Witnessed violence between adults in the home | 19305 | 91871 | 21.01 | 0.540 | 0.711 | -0.042 |
| Physical abuse | 23204 | 91871 | 25.26 | 0.524 | 0.713 | -0.040 |
| Emotional abuse | 41199 | 91871 | 44.84 | 0.530 | 0.711 | -0.042 |
| Sexual abuse | 23071 | 91871 | 25.11 | 0.345 | 0.746 | -0.007 |

Note: Internal consistency analysis of the derived eight-domain Adverse Childhood Experiences (ACE) burden score among participants with sufficient information to classify all eight ACE domains (N = 91,871). The overall Kuder–Richardson Formula 20 (KR-20) coefficient was **0.753**. For each ACE domain, the table reports the number and percentage of participants with a positive classification, the corrected item-total correlation, the KR-20 coefficient following deletion of the domain, and the corresponding change in KR-20 (ΔKR-20) relative to the full eight-domain ACE burden score.

**Supplementary Table S10.** Higher eight-domain ACE burden remained associated with all predefined outcomes after demographic adjustment.

| Outcome Domain | Outcome | N | Cases | Prevalence, % | Adjusted OR (95% CI) | p-value |
|---|---|---|---|---|---|---|
| Clinical | PTSD | 91871 | 2699 | 2.9 | 1.38 (1.36-1.41) | <0.001 |
| SDOH | Food insecurity | 84386 | 9728 | 11.5 | 1.37 (1.35-1.38) | <0.001 |
| Clinical | Bipolar disorder | 91871 | 1657 | 1.8 | 1.34 (1.31-1.36) | <0.001 |
| Clinical | Suicidal ideation/self-harm | 91871 | 861 | 0.9 | 1.31 (1.27-1.35) | <0.001 |
| Clinical | Substance use disorder | 91871 | 3203 | 3.5 | 1.26 (1.24-1.28) | <0.001 |
| SDOH | Social isolation | 82777 | 25368 | 30.6 | 1.22 (1.21-1.23) | <0.001 |
| SDOH | Unsafe neighborhood | 82782 | 5595 | 6.8 | 1.20 (1.18-1.22) | <0.001 |
| Clinical | Major depressive disorder | 91871 | 13953 | 15.2 | 1.16 (1.15-1.17) | <0.001 |
| Clinical | Generalized anxiety disorder | 91871 | 6066 | 6.6 | 1.12 (1.11-1.14) | <0.001 |
| Clinical | Sleep disorder | 91871 | 18388 | 20.0 | 1.07 (1.06-1.08) | <0.001 |
| Clinical | Obesity | 91871 | 15954 | 17.4 | 1.07 (1.06-1.08) | <0.001 |
| Clinical | Chronic pain | 91871 | 18706 | 20.4 | 1.06 (1.05-1.06) | <0.001 |

Note: Adjusted logistic regression analyses evaluating associations between the continuous eight-domain Adverse Childhood Experiences (ACE) burden score and predefined clinical and social determinant of health (SDOH) outcomes among participants with sufficient information to classify all eight ACE domains. Separate models were fit for each outcome, with the continuous ACE burden score (0–8) modeled as the primary exposure. Models were adjusted for age group, gender, race, and ethnicity. Results are presented as adjusted odds ratios (ORs) with 95% confidence intervals (CIs), representing the change in odds associated with each one-point increase in eight-domain ACE burden score.

**Supplementary Table S11.** Higher eight-domain ACE burden was associated with all predefined outcomes in unadjusted analyses.

| Outcome Domain | Outcome | N | Cases | Prevalence, % | Unadjusted OR (95% CI) | p-value |
|---|---|---|---|---|---|---|
| SDOH | Food insecurity | 84386 | 9728 | 11.5 | 1.47 (1.46-1.48) | <0.001 |
| Clinical | PTSD | 91871 | 2699 | 2.9 | 1.40 (1.38-1.42) | <0.001 |
| Clinical | Bipolar disorder | 91871 | 1657 | 1.8 | 1.38 (1.36-1.41) | <0.001 |

| | | | | | | |
|---|---|---|---|---|---|---|
| Clinical | Suicidal ideation/self-harm | 91871 | 861 | 0.9 | 1.35 (1.32-1.39) | <0.001 |
| SDOH | Social isolation | 82777 | 25368 | 30.6 | 1.26 (1.25-1.27) | <0.001 |
| SDOH | Unsafe neighborhood | 82782 | 5595 | 6.8 | 1.23 (1.22-1.25) | <0.001 |
| Clinical | Substance use disorder | 91871 | 3203 | 3.5 | 1.23 (1.21-1.25) | <0.001 |
| Clinical | Major depressive disorder | 91871 | 13953 | 15.2 | 1.17 (1.16-1.17) | <0.001 |
| Clinical | Generalized anxiety disorder | 91871 | 6066 | 6.6 | 1.16 (1.14-1.17) | <0.001 |
| Clinical | Obesity | 91871 | 15954 | 17.4 | 1.05 (1.05-1.06) | <0.001 |
| Clinical | Sleep disorder | 91871 | 18388 | 20.0 | 1.02 (1.02-1.03) | <0.001 |
| Clinical | Chronic pain | 91871 | 18706 | 20.4 | 1.02 (1.01-1.03) | <0.001 |

Note: Unadjusted logistic regression analyses evaluating associations between the continuous eight-domain Adverse Childhood Experiences (ACE) burden score and predefined clinical and social determinant of health (SDOH) outcomes among participants with sufficient information to classify all eight ACE domains. Separate models were fit for each outcome, with the continuous ACE burden score (0–8) modeled as the primary exposure. Results are presented as unadjusted odds ratios (ORs) with 95% confidence intervals (CIs), representing the change in odds associated with each one-point increase in eight-domain ACE burden score.

**Supplementary Table S12.** Outcome prevalence generally increased across eight-domain ACE burden scores.

| Outcome Domain | Outcome | ACE Burden Score | People, n | Cases, n | Prevalence, % |
|---|---|---|---|---|---|
| Clinical | Bipolar disorder | 0 | 26218 | 140 | 0.53 |
| Clinical | Bipolar disorder | 1 | 19090 | 178 | 0.93 |
| Clinical | Bipolar disorder | 2 | 13735 | 231 | 1.68 |
| Clinical | Bipolar disorder | 3 | 10674 | 231 | 2.16 |
| Clinical | Bipolar disorder | 4 | 8206 | 215 | 2.62 |
| Clinical | Bipolar disorder | 5 | 5906 | 225 | 3.81 |
| Clinical | Bipolar disorder | 6 | 4232 | 190 | 4.49 |
| Clinical | Bipolar disorder | 7 | 2677 | 157 | 5.86 |
| Clinical | Bipolar disorder | 8 | 1133 | 90 | 7.94 |
| Clinical | Chronic pain | 0 | 26218 | 5196 | 19.82 |
| Clinical | Chronic pain | 1 | 19090 | 3791 | 19.86 |
| Clinical | Chronic pain | 2 | 13735 | 2777 | 20.22 |
| Clinical | Chronic pain | 3 | 10674 | 2169 | 20.32 |
| Clinical | Chronic pain | 4 | 8206 | 1732 | 21.11 |
| Clinical | Chronic pain | 5 | 5906 | 1296 | 21.94 |

| | | | | | |
|---|---|---|---|---|---|
| Clinical | Chronic pain | 6 | 4232 | 907 | 21.43 |
| Clinical | Chronic pain | 7 | 2677 | 616 | 23.01 |
| Clinical | Chronic pain | 8 | 1133 | 222 | 19.59 |
| Clinical | Generalized anxiety disorder | 0 | 26218 | 1162 | 4.43 |
| Clinical | Generalized anxiety disorder | 1 | 19090 | 1039 | 5.44 |
| Clinical | Generalized anxiety disorder | 2 | 13735 | 924 | 6.73 |
| Clinical | Generalized anxiety disorder | 3 | 10674 | 798 | 7.48 |
| Clinical | Generalized anxiety disorder | 4 | 8206 | 734 | 8.94 |
| Clinical | Generalized anxiety disorder | 5 | 5906 | 553 | 9.36 |
| Clinical | Generalized anxiety disorder | 6 | 4232 | 422 | 9.97 |
| Clinical | Generalized anxiety disorder | 7 | 2677 | 287 | 10.72 |
| Clinical | Generalized anxiety disorder | 8 | 1133 | 147 | 12.97 |
| Clinical | Major depressive disorder | 0 | 26218 | 2624 | 10.01 |
| Clinical | Major depressive disorder | 1 | 19090 | 2538 | 13.29 |
| Clinical | Major depressive disorder | 2 | 13735 | 2091 | 15.22 |
| Clinical | Major depressive disorder | 3 | 10674 | 1873 | 17.55 |
| Clinical | Major depressive disorder | 4 | 8206 | 1637 | 19.95 |
| Clinical | Major depressive disorder | 5 | 5906 | 1308 | 22.15 |
| Clinical | Major depressive disorder | 6 | 4232 | 969 | 22.90 |
| Clinical | Major depressive disorder | 7 | 2677 | 656 | 24.51 |
| Clinical | Major depressive disorder | 8 | 1133 | 257 | 22.68 |
| Clinical | Obesity | 0 | 26218 | 4051 | 15.45 |
| Clinical | Obesity | 1 | 19090 | 3175 | 16.63 |
| Clinical | Obesity | 2 | 13735 | 2416 | 17.59 |
| Clinical | Obesity | 3 | 10674 | 1914 | 17.93 |
| Clinical | Obesity | 4 | 8206 | 1585 | 19.32 |
| Clinical | Obesity | 5 | 5906 | 1149 | 19.45 |
| Clinical | Obesity | 6 | 4232 | 885 | 20.91 |
| Clinical | Obesity | 7 | 2677 | 563 | 21.03 |
| Clinical | Obesity | 8 | 1133 | 216 | 19.06 |
| Clinical | PTSD | 0 | 26218 | 268 | 1.02 |
| Clinical | PTSD | 1 | 19090 | 300 | 1.57 |
| Clinical | PTSD | 2 | 13735 | 306 | 2.23 |
| Clinical | PTSD | 3 | 10674 | 321 | 3.01 |

| | | | | | |
|---|---|---|---|---|---|
| Clinical | PTSD | 4 | 8206 | 377 | 4.59 |
| Clinical | PTSD | 5 | 5906 | 368 | 6.23 |
| Clinical | PTSD | 6 | 4232 | 343 | 8.10 |
| Clinical | PTSD | 7 | 2677 | 276 | 10.31 |
| Clinical | PTSD | 8 | 1133 | 140 | 12.36 |
| Clinical | Sleep disorder | 0 | 26218 | 5004 | 19.09 |
| Clinical | Sleep disorder | 1 | 19090 | 3747 | 19.63 |
| Clinical | Sleep disorder | 2 | 13735 | 2741 | 19.96 |
| Clinical | Sleep disorder | 3 | 10674 | 2157 | 20.21 |
| Clinical | Sleep disorder | 4 | 8206 | 1707 | 20.80 |
| Clinical | Sleep disorder | 5 | 5906 | 1292 | 21.88 |
| Clinical | Sleep disorder | 6 | 4232 | 948 | 22.40 |
| Clinical | Sleep disorder | 7 | 2677 | 591 | 22.08 |
| Clinical | Sleep disorder | 8 | 1133 | 201 | 17.74 |
| Clinical | Substance use disorder | 0 | 26218 | 531 | 2.03 |
| Clinical | Substance use disorder | 1 | 19090 | 505 | 2.65 |
| Clinical | Substance use disorder | 2 | 13735 | 410 | 2.99 |
| Clinical | Substance use disorder | 3 | 10674 | 431 | 4.04 |
| Clinical | Substance use disorder | 4 | 8206 | 374 | 4.56 |
| Clinical | Substance use disorder | 5 | 5906 | 341 | 5.77 |
| Clinical | Substance use disorder | 6 | 4232 | 292 | 6.90 |
| Clinical | Substance use disorder | 7 | 2677 | 210 | 7.84 |
| Clinical | Substance use disorder | 8 | 1133 | 109 | 9.62 |
| Clinical | Suicidal ideation/self-harm | 0 | 26218 | 89 | 0.34 |
| Clinical | Suicidal ideation/self-harm | 1 | 19090 | 99 | 0.52 |
| Clinical | Suicidal ideation/self-harm | 2 | 13735 | 106 | 0.77 |
| Clinical | Suicidal ideation/self-harm | 3 | 10674 | 116 | 1.09 |
| Clinical | Suicidal ideation/self-harm | 4 | 8206 | 120 | 1.46 |
| Clinical | Suicidal ideation/self-harm | 5 | 5906 | 106 | 1.79 |
| Clinical | Suicidal ideation/self-harm | 6 | 4232 | 102 | 2.41 |
| Clinical | Suicidal ideation/self-harm | 7 | 2677 | 88 | 3.29 |
| Clinical | Suicidal ideation/self-harm | 8 | 1133 | 35 | 3.09 |
| SDOH | Food insecurity | 0 | 24203 | 1019 | 4.21 |
| SDOH | Food insecurity | 1 | 17570 | 1117 | 6.36 |

| | | | | | |
|---|---|---|---|---|---|
| SDOH | Food insecurity | 2 | 12660 | 1161 | 9.17 |
| SDOH | Food insecurity | 3 | 9799 | 1224 | 12.49 |
| SDOH | Food insecurity | 4 | 7537 | 1311 | 17.39 |
| SDOH | Food insecurity | 5 | 5393 | 1252 | 23.22 |
| SDOH | Food insecurity | 6 | 3827 | 1152 | 30.10 |
| SDOH | Food insecurity | 7 | 2394 | 957 | 39.97 |
| SDOH | Food insecurity | 8 | 1003 | 535 | 53.34 |
| SDOH | Social isolation | 0 | 23685 | 4518 | 19.08 |
| SDOH | Social isolation | 1 | 17203 | 4388 | 25.51 |
| SDOH | Social isolation | 2 | 12440 | 3882 | 31.21 |
| SDOH | Social isolation | 3 | 9608 | 3458 | 35.99 |
| SDOH | Social isolation | 4 | 7401 | 3001 | 40.55 |
| SDOH | Social isolation | 5 | 5312 | 2377 | 44.75 |
| SDOH | Social isolation | 6 | 3779 | 1833 | 48.50 |
| SDOH | Social isolation | 7 | 2355 | 1301 | 55.24 |
| SDOH | Social isolation | 8 | 994 | 610 | 61.37 |
| SDOH | Unsafe neighborhood | 0 | 23712 | 1033 | 4.36 |
| SDOH | Unsafe neighborhood | 1 | 17227 | 869 | 5.04 |
| SDOH | Unsafe neighborhood | 2 | 12423 | 738 | 5.94 |
| SDOH | Unsafe neighborhood | 3 | 9625 | 685 | 7.12 |
| SDOH | Unsafe neighborhood | 4 | 7405 | 658 | 8.89 |
| SDOH | Unsafe neighborhood | 5 | 5282 | 528 | 10.00 |
| SDOH | Unsafe neighborhood | 6 | 3765 | 460 | 12.22 |
| SDOH | Unsafe neighborhood | 7 | 2349 | 391 | 16.65 |
| SDOH | Unsafe neighborhood | 8 | 994 | 233 | 23.44 |

Note: Prevalence of predefined clinical and social determinant of health (SDOH) outcomes across eight-domain Adverse Childhood Experiences (ACE) burden scores among participants with sufficient information to classify all eight ACE domains. For each outcome and ACE burden score (0–8), the table reports the number of participants included in the analysis, the number of cases, and the corresponding prevalence. Sample sizes vary across outcomes because SDOH outcomes were available only for participants who completed the corresponding survey items, whereas clinical outcomes were evaluated using all participants with sufficient information to classify all eight ACE domains.

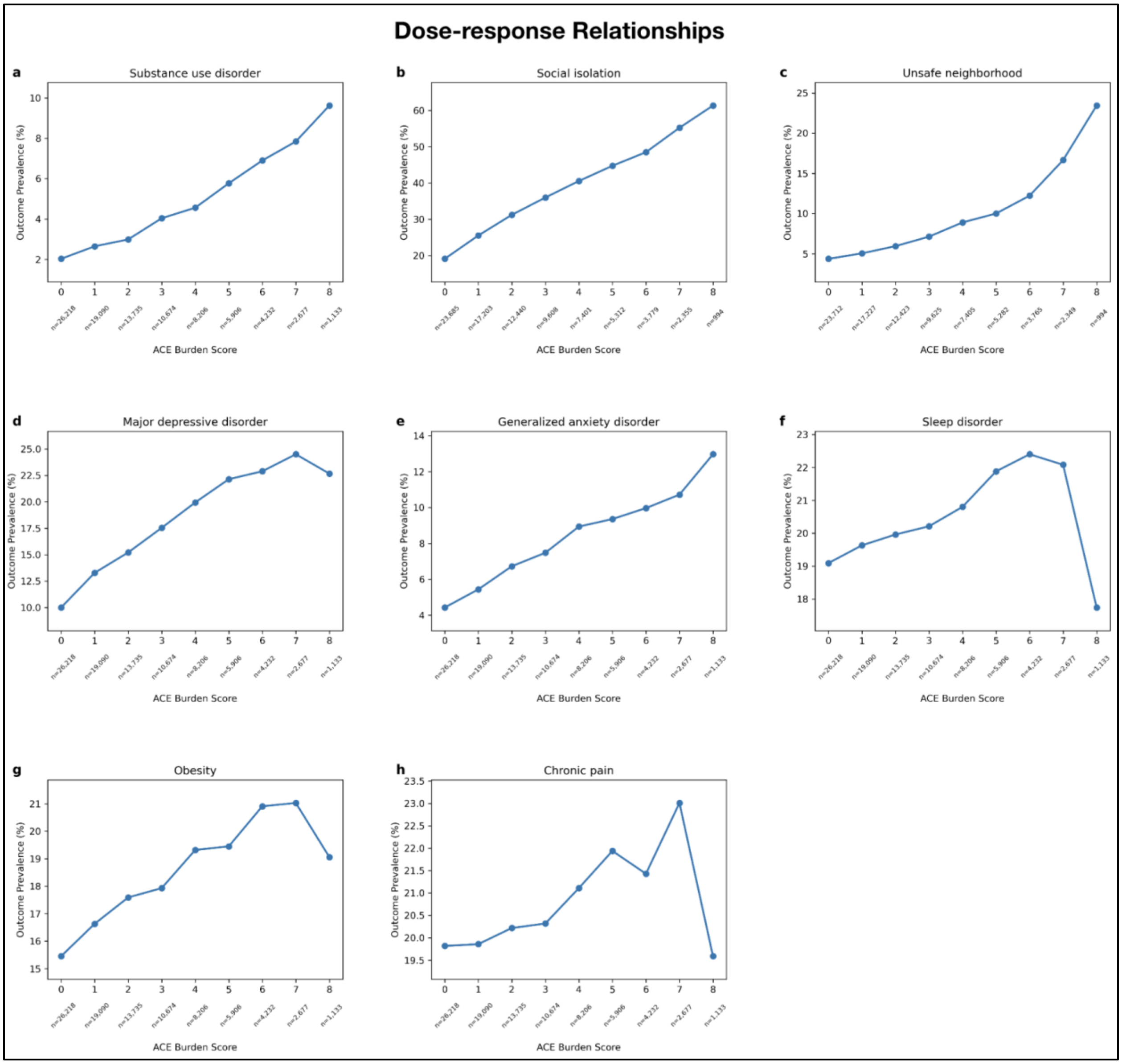


**Supplementary Figure S2.** The remaining predefined outcomes showed varying prevalence patterns across ACE burden scores. The eight outcomes shown are **(a)** substance use disorder, **(b)** social isolation, **(c)** unsafe neighborhood, **(d)** major depressive disorder, **(e)** generalized anxiety disorder, **(f)** sleep disorder, **(g)** obesity, and **(h)** chronic pain. Outcome prevalence was calculated for each ACE burden score (0–8) among participants with sufficient information to classify all eight ACE domains. Sample sizes for each ACE burden score category are displayed below the x-axis. Small deviations from a strictly increasing pattern at the highest ACE burden score reflect the relatively small number of participants with an ACE burden score of 8 (n = 1,133).

**Sensitivity Analysis.** Restricted cohort requiring ≥2 healthcare visits

**Supplementary Table S13.** Predefined outcome prevalence was estimated among 61,773 participants with at least two healthcare visits.

| Outcome | n with outcome | n total | % with outcome |
|---|---|---|---|
| MDD | 13277 | 61773 | 21.49 |
| PTSD | 2601 | 61773 | 4.21 |
| Bipolar disorder | 1529 | 61773 | 2.48 |
| Generalized anxiety disorder | 5781 | 61773 | 9.36 |
| Suicidal ideation/self-harm | 806 | 61773 | 1.30 |
| Substance use disorder | 3011 | 61773 | 4.87 |
| Chronic pain | 17983 | 61773 | 29.11 |
| Sleep disorder | 17580 | 61773 | 28.46 |
| Obesity | 15173 | 61773 | 24.56 |
| Food insecurity | 6226 | 57617 | 10.81 |
| Social isolation | 17014 | 56530 | 30.10 |
| Unsafe neighborhood | 3710 | 56623 | 6.55 |

Note: Prevalence of predefined clinical and social determinant of health (SDOH) outcomes among participants with sufficient information to classify all eight ACE domains who had at least two healthcare visits recorded in the All of Us electronic health record (N = 61,773). This restricted cohort was used as a sensitivity analysis to evaluate the robustness of the primary findings after limiting analyses to participants with greater longitudinal clinical observation. For each outcome, the table reports the number of participants with the outcome, the total number of participants included in the analysis, and the corresponding outcome prevalence. Sample sizes vary across outcomes because SDOH outcomes were available only for participants who completed the corresponding survey items.

**Supplementary Table S14.** Associations persisted after restriction to participants with at least two healthcare visits.

| Outcome Domain | Outcome | Model | N | Cases | Prevalence, % | OR (95% CI) | p-value |
|---|---|---|---|---|---|---|---|
| Clinical | MDD | Unadjusted | 61773 | 13277 | 21.5 | 1.20 (1.19–1.21) | <0.001 |
| Clinical | MDD | Adjusted | 61773 | 13277 | 21.5 | 1.18 (1.17–1.19) | <0.001 |
| Clinical | PTSD | Unadjusted | 61773 | 2601 | 4.2 | 1.43 (1.40–1.45) | <0.001 |
| Clinical | PTSD | Adjusted | 61773 | 2601 | 4.2 | 1.40 (1.37–1.42) | <0.001 |
| Clinical | Bipolar disorder | Unadjusted | 61773 | 1529 | 2.5 | 1.41 (1.38–1.44) | <0.001 |

| | | | | | | | |
|---|---|---|---|---|---|---|---|
| Clinical | Bipolar disorder | Adjusted | 61773 | 1529 | 2.5 | 1.35 (1.32–1.38) | <0.001 |
| Clinical | Generalized anxiety disorder | Unadjusted | 61773 | 5781 | 9.4 | 1.18 (1.16–1.19) | <0.001 |
| Clinical | Generalized anxiety disorder | Adjusted | 61773 | 5781 | 9.4 | 1.13 (1.12–1.15) | <0.001 |
| Clinical | Suicidal ideation/self-harm | Unadjusted | 61773 | 806 | 1.3 | 1.37 (1.33–1.41) | <0.001 |
| Clinical | Suicidal ideation/self-harm | Adjusted | 61773 | 806 | 1.3 | 1.32 (1.28–1.36) | <0.001 |
| Clinical | Substance use disorder | Unadjusted | 61773 | 3011 | 4.9 | 1.25 (1.23–1.27) | <0.001 |
| Clinical | Substance use disorder | Adjusted | 61773 | 3011 | 4.9 | 1.27 (1.25–1.29) | <0.001 |
| Clinical | Chronic pain | Unadjusted | 61773 | 17983 | 29.1 | 1.04 (1.03–1.05) | <0.001 |
| Clinical | Chronic pain | Adjusted | 61773 | 17983 | 29.1 | 1.07 (1.06–1.08) | <0.001 |
| Clinical | Sleep disorder | Unadjusted | 61773 | 17580 | 28.5 | 1.04 (1.03–1.05) | <0.001 |
| Clinical | Sleep disorder | Adjusted | 61773 | 17580 | 28.5 | 1.08 (1.07–1.09) | <0.001 |
| Clinical | Obesity | Unadjusted | 61773 | 15173 | 24.6 | 1.07 (1.06–1.08) | <0.001 |
| Clinical | Obesity | Adjusted | 61773 | 15173 | 24.6 | 1.08 (1.07–1.09) | <0.001 |
| SDOH | Food insecurity | Unadjusted | 57617 | 6226 | 10.8 | 1.47 (1.45–1.49) | <0.001 |
| SDOH | Food insecurity | Adjusted | 57617 | 6226 | 10.8 | 1.36 (1.35–1.38) | <0.001 |
| SDOH | Social isolation | Unadjusted | 56530 | 17014 | 30.1 | 1.27 (1.26–1.28) | <0.001 |
| SDOH | Social isolation | Adjusted | 56530 | 17014 | 30.1 | 1.22 (1.21–1.24) | <0.001 |
| SDOH | Unsafe neighborhood | Unadjusted | 56623 | 3710 | 6.6 | 1.23 (1.22–1.25) | <0.001 |
| SDOH | Unsafe neighborhood | Adjusted | 56623 | 3710 | 6.6 | 1.20 (1.18–1.22) | <0.001 |

Note: Unadjusted and adjusted logistic regression analyses evaluating associations between the continuous eight-domain Adverse Childhood Experiences (ACE) burden score and predefined clinical and social determinant of health (SDOH) outcomes among participants with sufficient information to classify all eight ACE domains who had at least two healthcare visits recorded in the All of Us electronic health record (N = 61,773). Odds ratios (ORs) represent the change in odds associated with each one-point increase in eight-domain ACE burden score. Adjusted models included age group, gender, race, and ethnicity as covariates. For each outcome, the table reports the number of participants included in the analysis, the number of cases, outcome prevalence, ORs with 95% confidence intervals (CIs), and corresponding p-values. Sample sizes vary across outcomes because SDOH outcomes were available only for participants who completed the corresponding survey items.

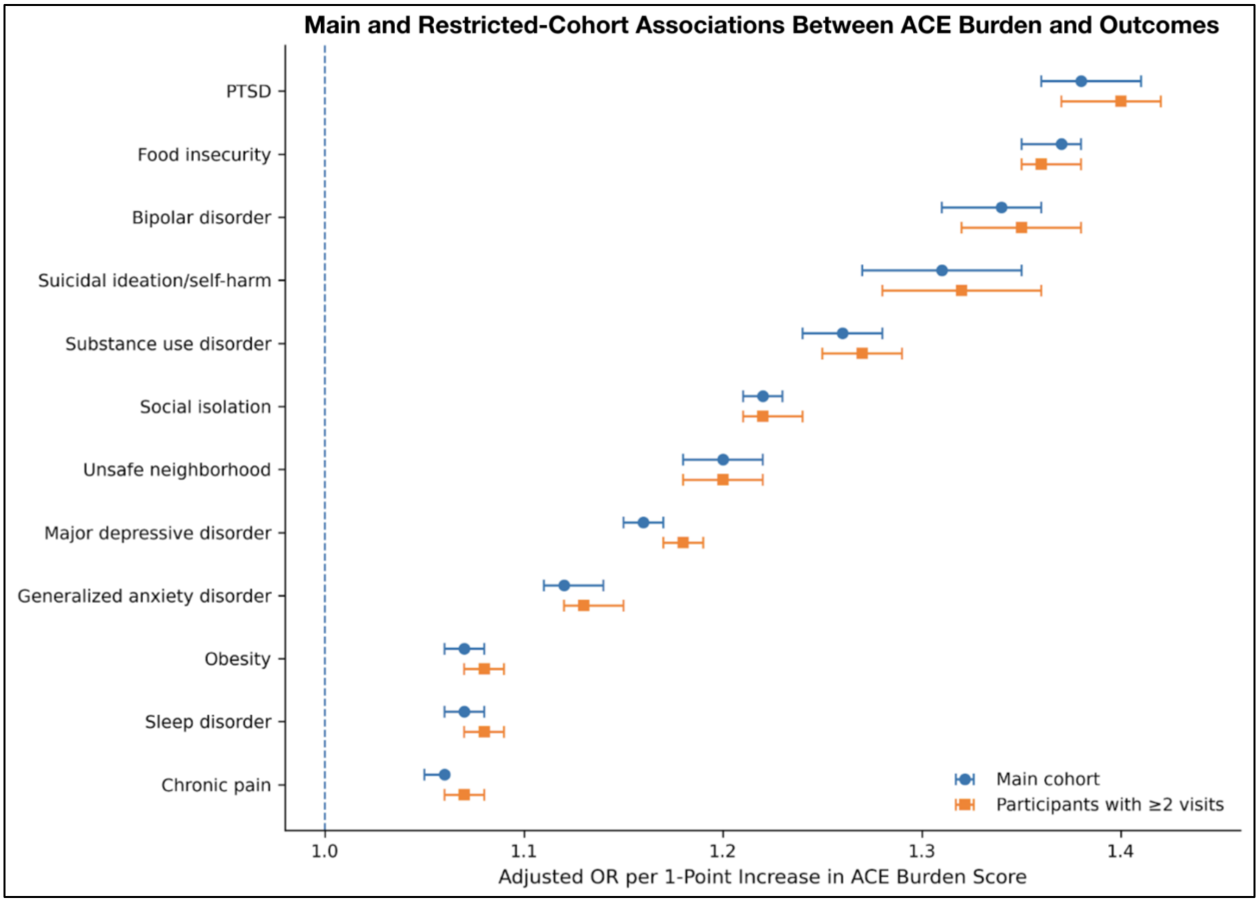


**Supplementary Figure S3.** Adjusted associations were similar after restriction to participants with at least two healthcare visits. Forest plot comparing adjusted odds ratios (ORs) and 95% confidence intervals (CIs) from the primary analysis with those from the sensitivity analysis restricted to participants with sufficient information to classify all eight ACE domains who had at least two healthcare visits recorded in the All of Us electronic health record. Odds ratios represent the change in odds associated with each one-point increase in the continuous eight-domain Adverse Childhood Experiences (ACE) burden score. All models were adjusted for age group, gender, race, and ethnicity.
